# Response strategies for COVID-19 epidemics in African settings: a mathematical modelling study

**DOI:** 10.1101/2020.04.27.20081711

**Authors:** Kevin van Zandvoort, Christopher I Jarvis, Carl A B Pearson, Nicholas G Davies, CMMID COVID-19 working group, Timothy W Russell, Adam J Kucharski, Mark Jit, Stefan Flasche, Rosalind M Eggo, Francesco Checchi

**Affiliations:** Centre for Mathematical Modelling of Infectious Diseases, Department of Infectious Disease Epidemiology, London School of Hygiene and Tropical Medicine, Keppel Street, WC1E 7HT London, UK; Health in Humanitarian Crises Centre, Department of Infectious Disease Epidemiology, London School of Hygiene & Tropical Medicine, Keppel Street, WC1E 7HT; South African Centre for Epidemiological Modelling and Analysis, Stellenbosch University, Stellenbosch, Republic of South Africa

**Keywords:** COVID-19, SARS-CoV-2, Coronavirus, Africa, Low-income, Control, Response, Mathematical model

## Abstract

**Background:** The health impact of COVID-19 may differ in African settings as compared to countries in Europe or China due to demographic, epidemiological, environmental and socio-economic factors. We evaluated strategies to reduce SARS-CoV-2 burden in African countries, so as to support decisions that balance minimising mortality, protecting health services and safeguarding livelihoods.

**Methods:** We used a Susceptible-Exposed-Infectious-Recovered mathematical model, stratified by age, to predict the evolution of COVID-19 epidemics in three countries representing a range of age distributions in Africa (from oldest to youngest average age: Mauritius, Nigeria and Niger), under various effectiveness assumptions for combinations of different non-pharmaceutical interventions: self-isolation of symptomatic people, physical distancing, and ‘shielding’ (physical isolation) of the high-risk population. We adapted model parameters to better represent uncertainty about what might be expected in African populations, in particular by shifting the distribution of severity risk towards younger ages and increasing the case-fatality ratio.

**Results:** We predicted median clinical attack rates over the first 12 months of 17% (Niger) to 39% (Mauritius), peaking at 2–4 months, if epidemics were unmitigated. Self-isolation while symptomatic had a maximum impact of about 30% on reducing severe cases, while the impact of physical distancing varied widely depending on percent contact reduction and *R_0_*. The effect of shielding high-risk people, e.g. by rehousing them in physical isolation, was sensitive mainly to residual contact with low-risk people, and to a lesser extent to contact among shielded individuals. Response strategies incorporating self-isolation of symptomatic individuals, moderate physical distancing and high uptake of shielding reduced predicted peak bed demand by 46% to 54% and mortality by 60% to 75%. Lockdowns delayed epidemics by about 3 months. Estimates were sensitive to differences in age-specific social mixing patterns, as published in the literature.

**Discussion:** In African settings, as elsewhere, current evidence suggests large COVID-19 epidemics are expected. However, African countries have fewer means to suppress transmission and manage cases. We found that self-isolation of symptomatic persons and general physical distancing are unlikely to avert very large epidemics, unless distancing takes the form of stringent lockdown measures. However, both interventions help to mitigate the epidemic. Shielding of high-risk individuals can reduce health service demand and, even more markedly, mortality if it features high uptake and low contact of shielded and unshielded people, with no increase in contact among shielded people. Strategies combining self-isolation, moderate physical distancing and shielding will probably achieve substantial reductions in mortality in African countries. Temporary lockdowns, where socioeconomically acceptable, can help gain crucial time for planning and expanding health service capacity.

## Introduction

The COVID-19 pandemic has not only led to increased mortality, but has also resulted in widespread socio-economic disruption and is severely testing affected countries’ health service capacity^1–3^. However, to date, its effects have mainly been observed in countries with relatively well-resourced health systems and the financial means to support economies during ‘lock-down’ periods. At the time of writing, only two African countries, Lesotho and Comoros, had not reported any confirmed SARS-CoV-2 infections^2^. In these and other low-income settings, two factors (younger age distributions and, potentially, warmer temperatures^4,5^) may help to attenuate the pandemic’s severity. However, other factors may plausibly combine to worsen its impact: these include demography (larger household sizes and more intergenerational mixing within households), environmental conditions (overcrowded urban settlements, inadequate water and sanitation), pre-existing disease burden (higher prevalence of undiagnosed or unmanaged non-communicable diseases, tuberculosis and, if confirmed to be risk factors for COVID-19 severity, HIV and undernutrition), and, critically, a very low baseline of and access to hospitalisation capacity, particularly intensive and sub-intensive care^6–10^. In several African countries, armed conflict, food insecurity and resulting forced displacement further worsen societal resilience^11–16^.

Options to manage COVID-19 in Africa may be limited. Sufficiently scaling up case management may simply be unfeasible for many countries as the requirements, particularly at the epidemic’s peak, may be many-fold greater than the baseline capacity. Even in scenarios where intense suppression measures are successfully implemented, it is plausible that the availability of beds, clinicians, ventilators and personal protective equipment would be critical limiting factors^17,18^. Suppressing the epidemic through ‘lock-down’ policies may delay transmission in the short-term, but their economic viability beyond a timeframe of weeks is questionable unless large economic rescue packages are made available by global financing actors and are concretely accessible to populations: indeed, lock-down measures and even less intense distancing restrictions could exacerbate poverty and undernutrition, compromise educational attainment and undo improvements in access to health interventions achieved over the past decades^19,20^.

To help inform COVID-19 response strategies for African settings, we undertook a mathematical modelling study. We explored the possible effect on hospitalisation requirements and mortality of interventions considered to date in high-income settings, including self-isolation of symptomatic persons, general distancing (reduction of overall contacts) outside the household and more intensive lock-down measures. We also quantified the potential of an alternative option we refer to as ‘shielding’, whereby people at high risk of COVID-19 severe disease are specifically protected through a variety of community-led arrangements, such as neighbourhood-level house swaps, to create ‘green zones’ wherein high-risk residents are physically isolated for an extended period, but supported to live safely and with dignity: epidemiologically, this option seeks to reduce transmission within the high-risk groups that may otherwise contribute a large amount of hospitalisation and mortality.

## Methods

### Model structure

We adapted a previously developed discrete-time Susceptible-Exposed-Infectious-Recovered (SEIR) compartmental model, stratified by age group and disease status (asymptomatic, pre-symptomatic, and symptomatic)^21^. Detail is provided in the Supplementary Material. In brief, the model progresses a population through time based on assumed age-dependent contact of susceptible with infectious individuals. After their infectious period, all individuals are assumed to be immune until the end of the simulation (Recovered compartment). The model is stratified into 16 age groups, with people under 75 years stratified into 5-year age-bands and one additional stratum for 75+ years. The model population is closed, with no births or ageing, and deaths remain in the Recovered compartment.

### Transmissibility assumptions

We adopted epidemiological parameter values (serial interval, infectiousness by symptom stage, and incubation, infectiousness and symptomatic periods) used by Davies et al.^21^ (see Supplementary Material). We assumed an age-dependent probability of developing clinical symptoms.^22^ Asymptomatic individuals were assumed to be half as infectious as symptomatic individuals. Clinical progression of symptomatic cases to severe disease is assumed not to affect their infectiousness.

To represent the full range of age structures in Africa, we ranked countries according to their mean age, and selected countries with the youngest (Niger), oldest (Mauritius), and median (Nigeria) mean age as case studies. Key country-specific data inputs were age-specific population sizes, sourced from United Nations World Population Prospects estimates^23^ and age- and setting-specific social contact matrices. As no empirical data on age-specific social mixing patterns were available for these three countries, we used previously published synthetic contact matrices, namely projections of a European multi-country contact pattern study adjusted to individual countries based on national demographic and socio-economic characteristics^24^. Contact data were stratified according to whether the contact was within or outside the household.

To account for uncertainty regarding the transmissibility of SARS-CoV-2 in Africa, we implemented the model stochastically and sampled the basic reproduction number *R*_0_, from a normal distribution with mean 2.6 and standard deviation of 0.5^25^. We implemented different *R*_0_ estimates by scaling the probability of transmission per contact with an infectious person in accordance with the ratio between the target *R*_0_ and the dominant eigenvalue of the Next-Generation Matrix (Supplementary Material).

### Case severity assumptions

In high-income countries, the severity of SARS-CoV-2 infections has been shown to increase with age and prevalence of various comorbidities^26–28^. In practice, individual comorbidities co-occur (e.g. diabetes and hypertension), and co- and multimorbidity increase with age^10^. To simplify assumptions, we took age as a single predictor of severity, applying current evidence on its association with risk of symptomatic disease, severe disease (i.e. requiring hospitalisation), and critical disease (need for intensive care).

However, in African and low-income countries, an average person’s underlying vulnerability may correspond to that of an individual with greater chronological age in a high-income setting due to life-course effects including malnutrition, infections and often unmanaged non-communicable diseases. It is not yet known how this might affect COVID-19 severity^29^, although Global Burden of Disease data show strong associations between income level and the severity of other respiratory infections, particularly in younger age groups.^29^ To account for this, we shifted age-specific severity risks (probability of becoming a severe case and critical case) towards younger age by 10 years. To explore the effect of increased vulnerability and lack of access to healthcare, we also multiplied current estimates of age-specific case-fatality ratios (CFR; from China and the Diamond Princess cruise ship outbreak) of severe, and critical cases by a factor of 1.5 (used for our main analysis; see Supplementary Material). We did not make assumptions about the proportion of cases that would receive appropriate treatment: the CFR multiplier factor attempts to capture worse prognosis under limited or no treatment.

### Response interventions

The range of response interventions explored, alone or in combination, are outlined in Table 1. We assumed these would be applied at the country level.

Self-isolation was implemented as a reduction in transmissibility of infected people during their symptomatic period, equivalent to a reduction in their contacts. We did not account for additional quarantine of other members in the same household.

General physical distancing was implemented as a reduction in all contacts outside the household. We assumed no change in transmission within the household.

Shielding was implemented by stratifying the population into one shielded and one unshielded compartment. In the presence of shielding, mixing between the shielded and unshielded population would reduce by some degree, while mixing within the shielded population may remain the same, decrease (to zero if people are shielded individually) or increase if shielded people are resettled in overcrowded housing. While in the intervention’s practical application people should be shielded on the basis of age and/or known comorbidities, for this model we assumed more simplistically that varying proportions of the population aged 60 and above would be shielded.

**Table 1.** Summary of response interventions explored in the study.

| Intervention | Description | Model implementation | Range explored |
| --- | --- | --- | --- |
| Self-isolation of symptomatic people | People with symptoms of possible COVID-19 isolate themselves in their home until symptom-free | Relative reduction in all social contacts among symptomatic people only, from the time they become symptomatic | 0-100% relative reduction in transmission |
| General physical distancing (including reduction of probability of transmission per contact) | Behaviour change, promotion of handwashing, varying degrees of curtailment of transport, social and work gatherings | Relative reduction in contacts outside of the household | 0-100% relative reduction in transmission (we assumed that 'lockdown' measures would correspond to an 80% reduction)<sup>24</sup> |
| Shielding of high-risk groups | Communities identify people who meet high-risk criteria for poor COVID-19 clinical outcomes and resettle them in a variety of shielded arrangements (either individual, e.g. a dedicated room within a house, or groups of various sizes in vacated / swapped houses, huts or other shelters). Contact is thereafter limited.<br>Within these shielded 'green zones', residents also adopt distancing measures if they are living together. | A proportion of people aged $\geq 60$ years old is 'shielded'<br>Contact between shielded people and unshielded is reduced<br>Contact among shielded people varies depending on the arrangement chosen | 60-100% of high-risk people are shielded<br>60-100% relative reduction in contact with non-shielded people<br>0-400% relative change in social contact among shielded people (0% represents individual shielding arrangements; 400% represents what might happen if people are grouped within overcrowded shielded housing) |

### Analysis outcomes

We compared outcomes under different intervention values and strategies (combinations of interventions) for each unique combination of sampled *R*_0_ and model seeds. We present median, 50% and 95% quantiles of differences in outcomes across all combinations, and show results stratified by *R*_0_ in the Supplementary Material.

For the impact of individual interventions, we considered severe cases as our primary outcome. For the impact of different strategies, we observed the total number of symptomatic cases, severe cases (those who require hospitalisation), critical cases (those who require intensive care, ICU), and deaths, and present the expected time until peak of the epidemic, including peak bed demand that would be required. We only show estimates for the first 12 months after introduction of the first case, as the evolution of the epidemic beyond this period is subject to considerable unknowns (e.g. availability of vaccines or therapeutics; virus mutation, persistence of natural immunity).

### Ethics

The study used only publicly available aggregate data and was thus not subject to ethical review.

## Results

### Epidemic trajectories in the absence of control

Simulations of an unmitigated epidemic in Niger resulted in a median of 4.1 million clinical cases during the first 12 months following introduction of the first case, 48.7 million in Nigeria and 490 000 in Mauritius (Table 2), with the most probable epidemic peaks after 3, 4 and 2 months respectively (Figure 1). We estimate some 39,000 deaths due to COVID-19 in Niger, 605,000 in Nigeria and 17,000 in Mauritius would occur over the same period, not accounting for indirect excess mortality due to health service or socioeconomic disruptions. However, there is considerable uncertainty associated with these median estimates of cases and deaths (Table 2), which are mainly the result of different *R*_0_ values considered. Large epidemics that peak early occur in scenarios where *R*_0_ is high, whereas epidemics with lower *R*_0_ will have a lower total and peak epidemic size, and will peak later (Supplemental Figure S9).

**Figure 1:**
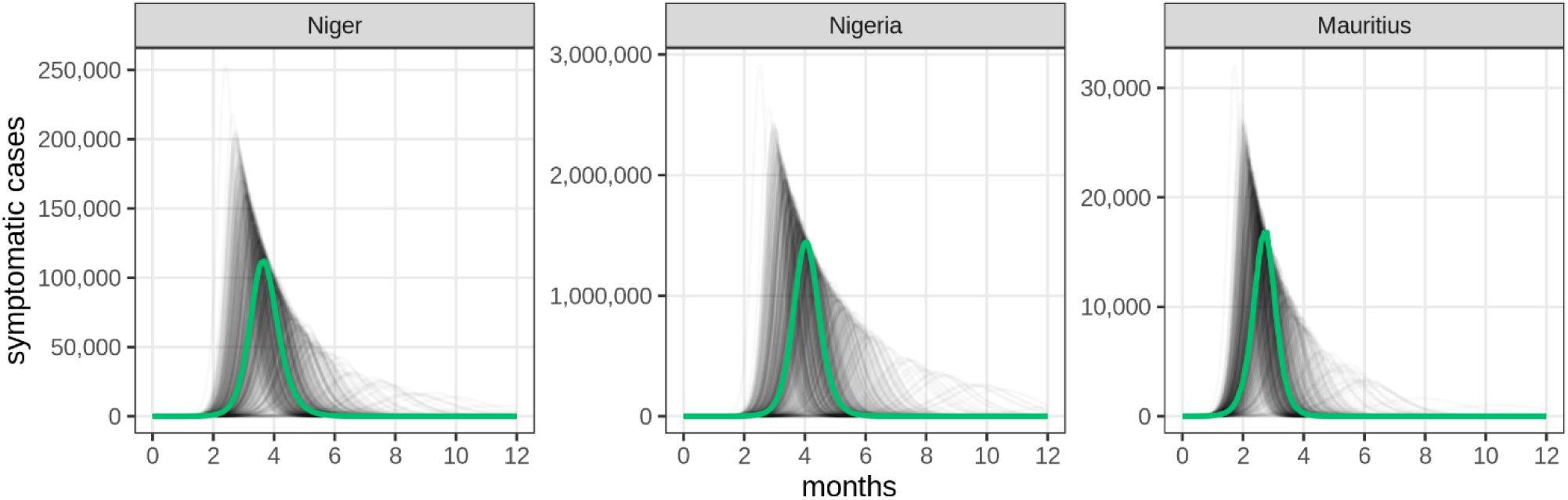
Projected incidence of symptomatic COVID-19 cases over time for simulations of an unmitigated epidemic, by country. The green line shows the run that was closest to the median total number of cases across all model runs. Grey lines show individual stochastic model runs, where *R*_0_ in each run was sampled from a normal distribution with mean 2.6 and standard deviation 0.5. Stratification of these runs by *R*_0_ are shown in the Supplementary Material.

### Effect of individual interventions

#### Self-isolation of symptomatic individuals

We estimate a reduction in severe cases during the first 12 months of the epidemic if symptomatic cases self-isolate throughout this period with varying levels of adherence (Figure 2A). Increasing adherence has a nearly linear relationship with the incidence of severe cases, but the maximum median impact is a 30% reduction (under an extreme scenario of 100% reduction in all contacts while having clinical symptoms). Although the probability of being a (symptomatic) case is modelled age-dependent, the impact of self-isolation was similar in all countries despite their different demographic distributions. Uncertainty intervals for Nigeria and Mauritius were wider, but mainly reflected differences in *R*_0_ between simulations (Supplemental Figure S10). In simulations where a low *R*_0_ was used, there was potential for a larger reduction in severe cases. The impact of *R*_0_ variability is smaller in Niger, where a higher proportion of transmission is due to subclinical infections due to its lower average age and the age-dependency of becoming a symptomatic case, as reflected by the clinical attack rates in Table 2.

**Table 2:** Projected impact of unmitigated COVID-19 epidemics during the first 12 months following introduction of cases, by country. All values represent the median and 95% lower and upper quantiles from 500 model runs. The symptomatic attack rate is calculated as the total number of symptomatic cases divided by the population. We show the months until the epidemic peak (defined as the day with the highest number of new cases) and present the peak daily number of deaths and hospital bed demand.

| Key outcome | Niger | Nigeria | Mauritius |
| --- | --- | --- | --- |
| Population size | 24,100,000 | 202,900,000 | 1,300,000 |
| Population aged 60+ | 4% | 5% | 18% |
| Symptomatic cases | 4,100,000<br>(1,800,000 to 5,200,000) | 48,700,000<br>(27,800,000 to 56,500,000) | 490,000<br>(314,000 to 546,000) |
| Severe cases | 106,000<br>(39,000 to 154,000) | 1,600,000<br>(734,000 to 2,100,000) | 44,000<br>(24,000 to 53,000) |
| Critical cases | 45,000<br>(17,000 to 66,000) | 699,000<br>(314,000 to 908,000) | 19,000<br>(10,000 to 23,000) |
| Symptomatic attack rate | 17% (8 to 22) | 24% (14 to 27) | 39% (25 to 43) |
| Deaths | 39,000<br>(14,000 to 56,000) | 605,000<br>(271,000 to 790,000) | 17,000<br>(9,000 to 21,000) |
| Deaths per 1000 person-years | 1.6 (0.6 to 2.3) | 2.9 (1.3 to 3.8) | 13.2 (7.1 to 16.2) |
| Epidemic peak (month) | 3 (3 to 7) | 4 (3 to 8) | 2 (2 to 5) |
| Peak deaths | 1500<br>(300 to 2800) | 25,600<br>(5700 to 43,200) | 800<br>(200 to 1200) |
| Peak demand for non-ICU beds | 23,200<br>(4400 to 43,200) | 391,000<br>(85,800 to 662,900) | 11,500<br>(3000 to 18,000) |
| Peak demand for ICU beds | 12,200<br>(2300 to 22,500) | 204,500<br>(45,300 to 343,900) | 6,000<br>(1600 to 9300) |

#### Population-wide physical distancing

Figure 2B shows the estimated impact of population-wide (i.e. not targeting any group) physical distancing, whereby all individuals reduce their contacts outside of the household to a certain degree, while contacts within the household remain unchanged. Across all three countries, reducing all contacts outside the household by an extreme of 100%, if sustained over the entire 12 months period, could result in a median reduction in severe cases by over 90%. However, this would largely delay rather than prevent severe cases, as insufficient levels of herd-immunity would develop, leading to a second wave of cases following the relaxation of measures. Relatively high reductions in contacts are needed for large impacts. Patterns were largely consistent across countries. Stratified results by *R*_0_ are shown in Supplemental Figure S10. Impact can vary substantially between settings with low and high *R*_0_, reflected by the wide intervals here. For instance, when reducing 40% of contacts outside of the household, reduction in severe cases in the first 12 months of the epidemic could be as low as 18% in scenarios with an *R*_0_ above 3, while it could be as high as 71% in scenarios with an *R*_0_ below 2.

**Figure 2:**
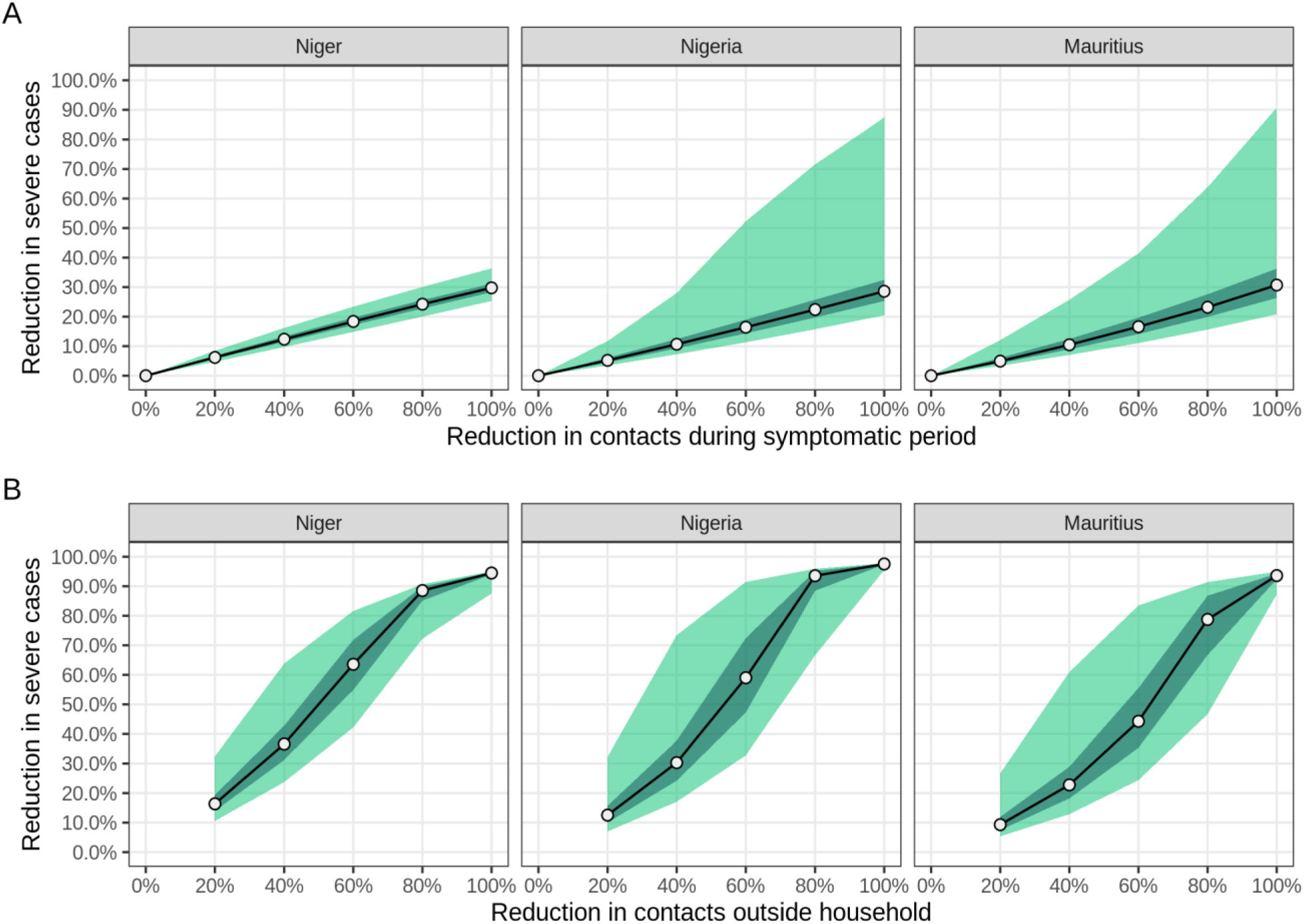
Estimated reduction in severe cases following A) self-isolation of symptomatic individuals and B) population-wide physical distancing. Medians (circles), 95% (light-green area) and 50% (dark green area) quantiles for the percentage reduction in severe cases during the first 12 months of the epidemic for different levels of compliance, for each country, across 500 model runs. Quantiles are calculated across all simulations representing different stochastic runs and using different *R*_0_ values. Estimates for reductions where no point is available are interpolated.

### Shielding of high-risk individuals

Shielding of high-risk individuals aims to reduce the number of severe cases among high-risk groups, and thereby in the overall population, while having a smaller effect on transmission in the population and thereby on the total number of cases. Figure 3 shows the reduction in the number of severe cases for different reductions in contacts between shielded and unshielded individuals, percentages of individuals shielded and changes in contact intensity within the shielded group, relative to baseline.

**Figure 3:**
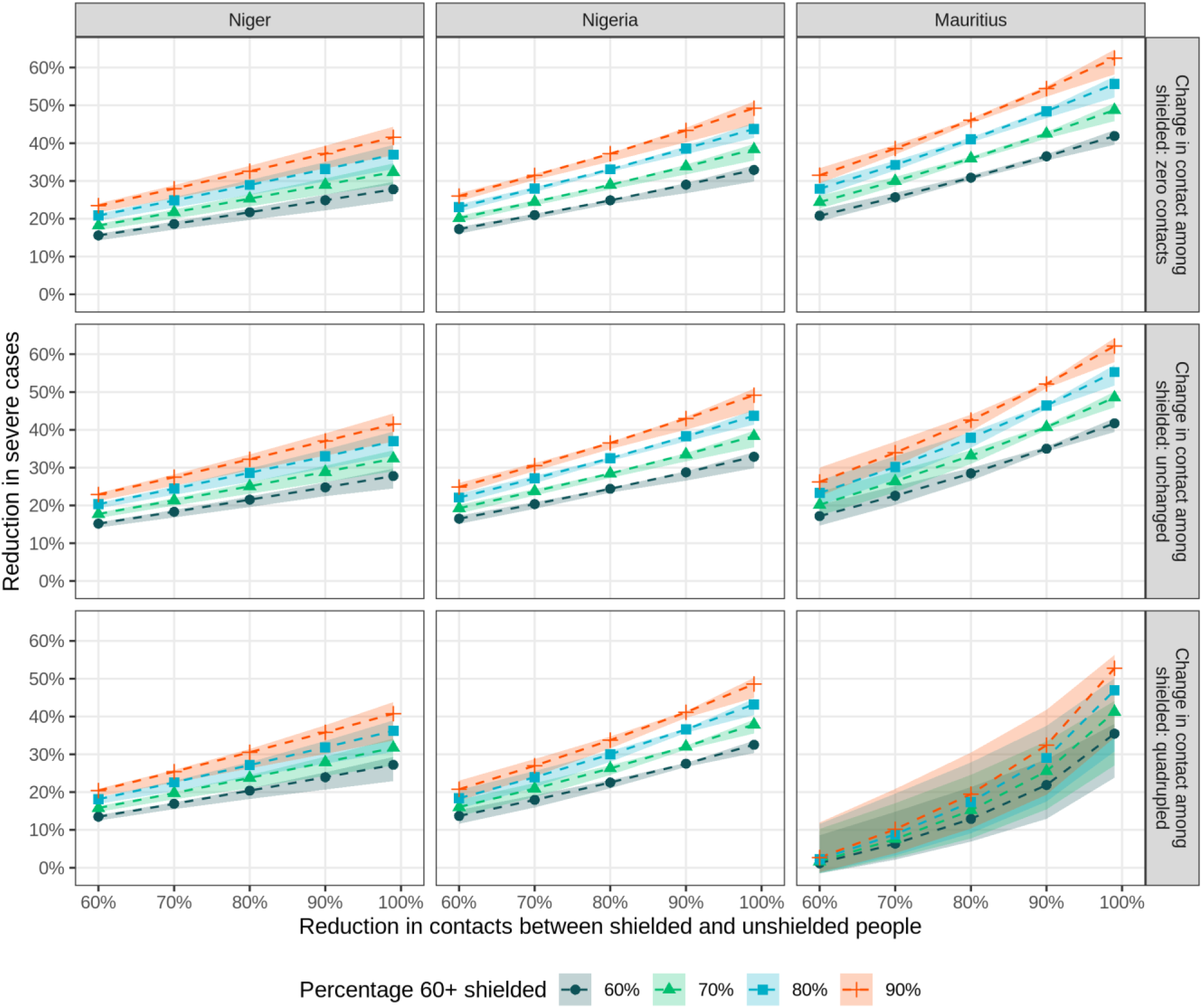
Estimated reduction in severe cases when shielding high-risk individuals, by country. Medians (dashed lines) and 95% quantiles (shaded areas) of the percentage reduction in severe cases during the first 12 months of the epidemic for different levels of reduction in contacts between shielded and unshielded people (x axis), different level of contacts among shielded people (facet rows), and for different percentages of people ≥ 60 years old shielded (see legend), for each country, across 500 model runs.

Across all countries, reductions in severe cases increased with the percentage ≥ 60 years old shielded, but the reduction in contacts between shielded and unshielded individuals was more influential, with ≥ 60% reduction in contacts required to achieve ≥ 10% reduction in severe cases. The degree of contact among shielded individuals appeared to be of lesser importance for Niger and Nigeria, with similar effect sizes regardless of whether the shielded individuals reduce their contact with one another to zero, remain at baseline, or quadruple it. This pattern does not hold for Mauritius, where a marked drop in the effect is seen when contact among shielded individuals quadruples: this is reflective of Mauritius having a larger elderly population, thereby contributing more to the overall proportion of severe cases in the population.

As shielding does not significantly affect transmission dynamics, estimates are similar across scenarios with low- and high *R*_0_ (Supplemental Figure S11). However, prediction intervals are wider in Mauritius, where a high proportion of the total population is shielded (8 - 14%).

### Impact of potential control strategies

#### Without lockdowns

We explored the impact of five different strategies and compared their impact to the unmitigated epidemic over the first 12 months after introduction of the first case. We assumed that self-isolation of symptomatic individuals, featuring 50% reduction in transmission, would be part of any strategy. We then added (i) 20% or (ii) 50% reduction in contacts outside the household through physical distancing, (iii) shielding of 80% of individuals aged 60 and older, with a reduction of 80% in contacts between the shielded and unshielded population and no change in contacts within the shielded population, (iv) a combination of shielding and 20% physical distancing, and (v) a combination of shielding with 50% physical distancing. Interventions are implemented when daily incidence reaches 1 case per 10,000 people and are assumed to be maintained for the remainder of the year.

Figure 4 shows the evolution of deaths depending on the strategy chosen. Evolution of bed demand under each scenario is given in Supplementary Figure S2. Table 3 shows the corresponding attack rate, total number of cases, severe cases, and critical cases, time of epidemic peak, and peak bed demand for severe and critical cases. All strategies yielded substantial but partial reductions in key health outcomes. Under all strategies, we estimate a high bed capacity needed in the three countries modelled.

**Figure 4:**
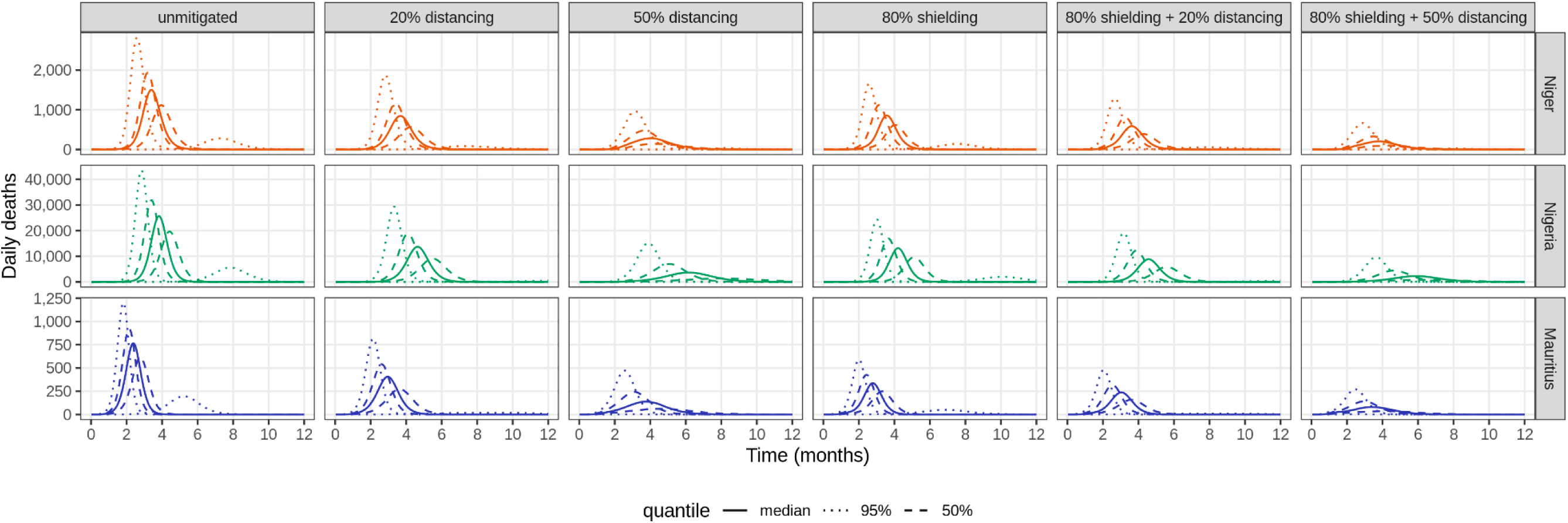
Estimated daily number of deaths during the first 12 months of the epidemic, under different strategies. Thick solid lines show the run which was closest to the median total number of deaths after 12 months across all model runs. Dashed lines are runs closest to the lower and upper 95% quantiles, while dotted lines are runs closest to the lower and upper 50% quantiles of total number of deaths, calculated over 500 model runs. Except for the unmitigated scenario, all scenarios assume 50% self-isolation during the symptomatic period of all clinical cases throughout the entire course of the epidemic. Other interventions start when daily incidence of symptomatic cases reaches 1 case per 10 000 people. Distancing strategies assume 20% or 50% reduction in all contacts outside of the household. Shielding strategies assume shielding of 80% of the population aged 60+, irrespective of underlying comorbidities, with an 80% reduction in contacts between the shielded and unshielded population, and no change in contacts within the shielded population. Estimates for bed demand over time are given in Supplementary Figure S2.

Whereas reducing transmission outside of the household by 20% would be more effective in reducing the total number of clinical cases than shielding, shielding could be as effective in reducing the total bed demand at the peak of the epidemic and total number of deaths as general physical distancing.

More substantial levels of physical distancing (50%) would lead to far greater effects in the first 12 months of the epidemic. However, this strategy, unlike shielding, reduces overall transmission and thus does not necessarily result in a resolution of the epidemic through herd immunity during this period; instead, this intervention would need to be sustained into the second year until herd-immunity is reached through either natural immunity or a vaccine, assuming immunity is long lived. Supplemental Figure S3 illustrates this phenomenon: when all interventions except self-isolation are lifted after 12 months, scenarios where physical distancing has reduced transmission outside the household by 50% in the first year may feature a second, albeit smaller peak during the second year if no further mitigation measures are taken; this further peak is absent under the other strategies. However, predictions beyond the first year may vary considerably depending on the longevity of SARS-CoV-2 immunity and availability of potential pharmaceutical interventions, and should be interpreted with caution.^4^

A combination of shielding and physical distancing would be most effective in reducing overall outcomes. Table 4 highlights the relative reduction in hospital bed demand for severe cases at the epidemic peak and total number of deaths in the first 12 months of the epidemic, under each scenario. Notably, shielding 80% of the high-risk population and reducing contact between the shielded and unshielded populations by 80% could be as effective in reducing severe outcomes as general physical distancing reducing transmission with 20%.

#### In combination with lockdowns

We also explored the impact a temporary lockdown could have on these same strategies. We assumed a two-month lockdown, triggered at incidence 1 per 10,000 person-days, during which, in addition to self-isolation as above, all contacts outside the household were reduced by 80%; the remainder of the year consisted of the five alternative strategies above.

Supplemental Figure S4 shows bed demand and deaths over time while Supplemental Table S3 shows key outcomes in the first 12 months under this lockdown scenario. A lockdown would delay, but not prevent, the epidemic peak in all countries (from 3 to 6–7 months in Niger, from 4 to 7–8 months in Nigeria, and from 2 to 5–6 months in Mauritius). However, it would not substantially affect total epidemic sizes or peak bed demand, compared to strategies without lockdowns.

#### Removing self-isolation of symptomatic individuals

As a sensitivity analysis, we explored how the impact of different scenarios would compare in the absence of self-isolation. Supplemental Figure S5 shows bed demand and deaths over time for each modelled strategy in the absence of self-isolation, while Supplemental Table S4 shows associated key outcomes.

Removing self-isolation substantially increased the number of cases and deaths in all scenarios. However, this relative difference was larger for scenarios with physical distancing only (11–15% higher mortality) than for scenarios with shielding only (6–10%). The resulting increased transmission brought the epidemic peak forward in all countries.

**Table 3:**
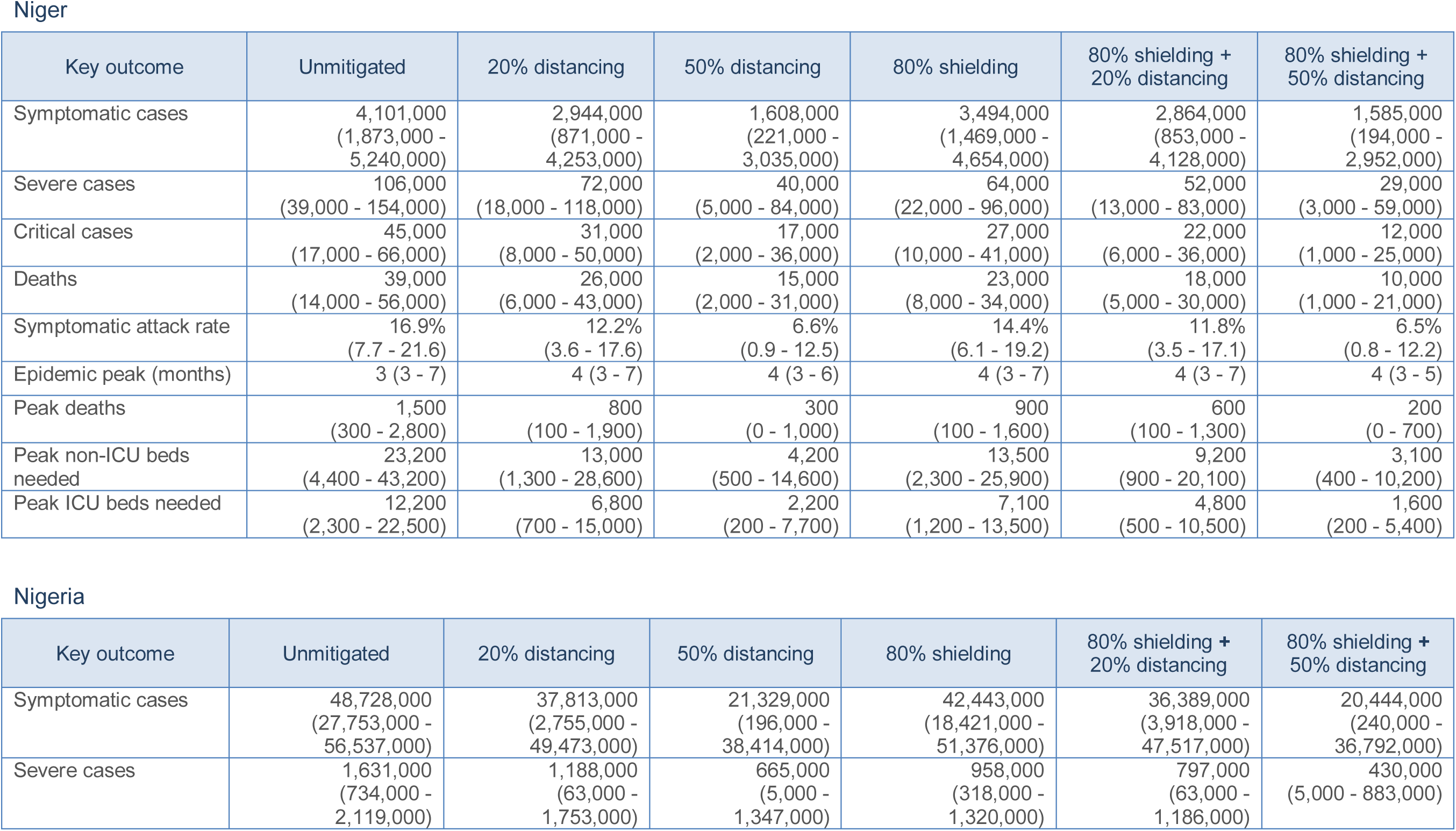
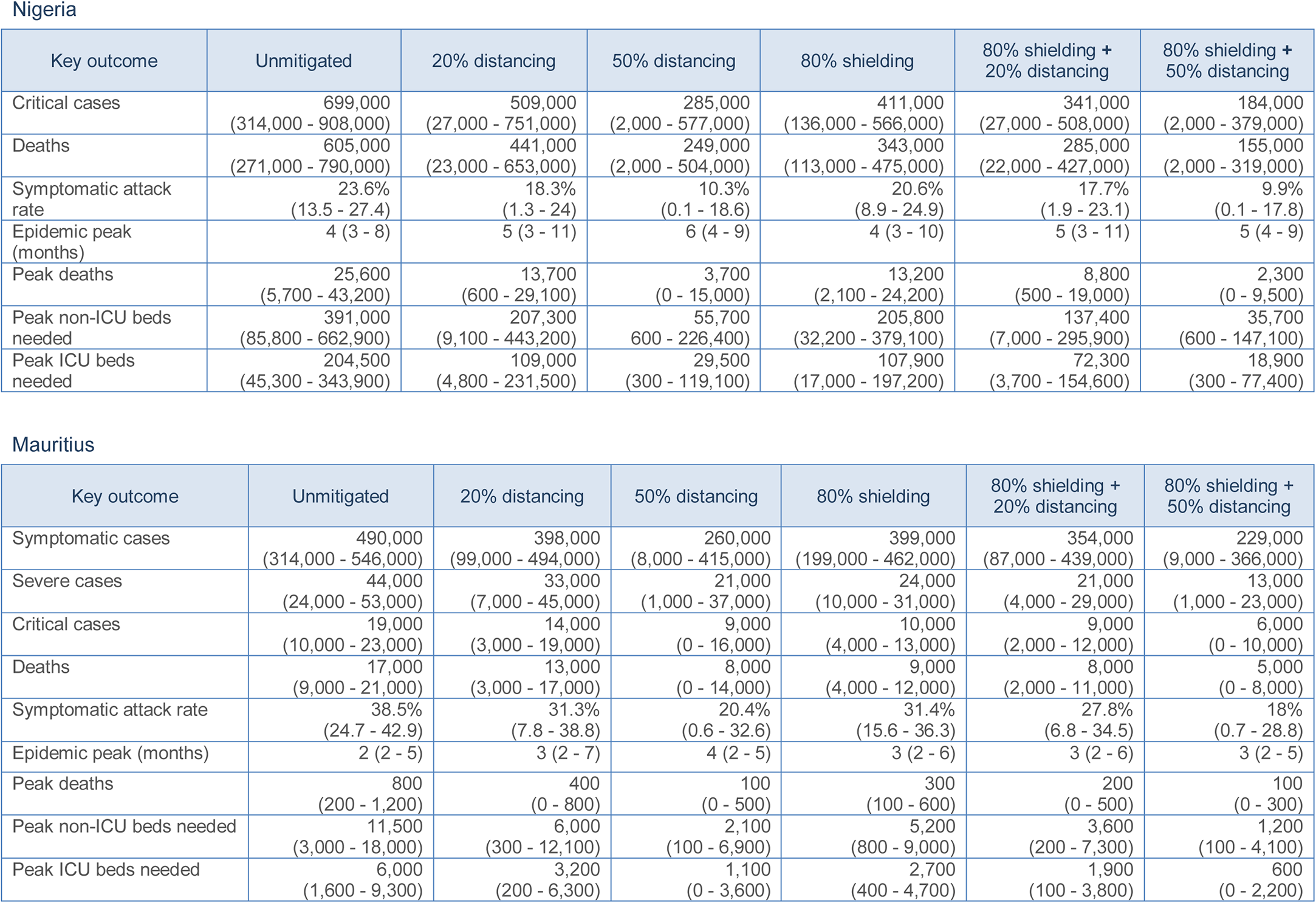
Key outcomes during the first 12 months of the epidemic under different model strategies, by country. Number of clinical, severe, and critical cases and deaths per 10,000 people predicted during the first 12 months of the epidemic, and number of deaths, non-ICU and ICU bed demand at epidemic peak in Niger, Nigeria, and Mauritius. Symptomatic attack rate is calculated as number of symptomatic cases over the total population size. We show estimates under different modelled strategies: (i) physical distancing with 20% reduction and (ii) 50% reduction in contacts outside of the household; (iii) shielding of 80% of the population over 60, with an 80% reduction in contacts between the shielded and unshielded population and no change in contacts within the shielded population; (iv) combined shielding and physical distancing with 20% and (v) 50% reduction in contacts outside of the household. In all modelled strategies, infected individuals decrease their contacts during their symptomatic infectious period by 50%.

**Table 4:**
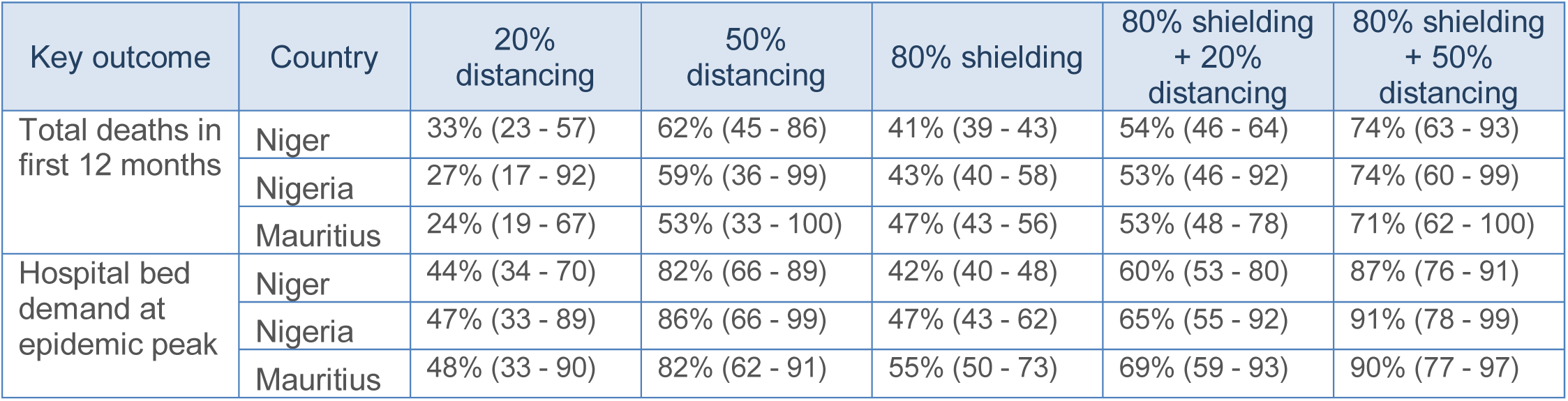
Relative reductions in two key outcomes during the first 12 months of the epidemic under different strategies. Table shows the relative reduction in the number of beds needed at the epidemic peak, and the total number of deaths in the first 12 months of the epidemic, compared to an unmitigated epidemic, under each strategy.

| Key outcome | Country | 20% distancing | 50% distancing | 80% shielding | 80% shielding + 20% distancing | 80% shielding + 50% distancing |
| --- | --- | --- | --- | --- | --- | --- |
| Total deaths in first 12 months | Niger | 33% (23 - 57) | 62% (45 - 86) | 41% (39 - 43) | 54% (46 - 64) | 74% (63 - 93) |
|  | Nigeria | 27% (17 - 92) | 59% (36 - 99) | 43% (40 - 58) | 53% (46 - 92) | 74% (60 - 99) |
|  | Mauritius | 24% (19 - 67) | 53% (33 - 100) | 47% (43 - 56) | 53% (48 - 78) | 71% (62 - 100) |
| Hospital bed demand at epidemic peak | Niger | 44% (34 - 70) | 82% (66 - 89) | 42% (40 - 48) | 60% (53 - 80) | 87% (76 - 91) |
|  | Nigeria | 47% (33 - 89) | 86% (66 - 99) | 47% (43 - 62) | 65% (55 - 92) | 91% (78 - 99) |
|  | Mauritius | 48% (33 - 90) | 82% (62 - 91) | 55% (50 - 73) | 69% (59 - 93) | 90% (77 - 97) |

#### Empirical contact matrices

To test the sensitivity of our results to this assumption, we replicated our analysis in three African countries for which empirical contact data are available: Kenya (rural Kilifi District), Uganda (rural to semi-urban Mbarara District), and Zimbabwe (the city of Bulawayo)^30–32^. To account for sampling error in these contact studies, we generated bootstrapped matrices for each of 500 model runs. Results are shown in the Supplementary Material. The impact of self-isolation and physical distancing was lower in these countries compared to our estimates for Niger, Nigeria, and Mauritius, though the impact of the shielding approach was similar or higher. We may therefore have underestimated the relative impact of shielding compared to the other interventions in our analysis, though impact of shielding was also lower in scenarios where contact within the shielded population increased, compared to the impact found in Niger, Nigeria, and Mauritius.

## Discussion

### Main findings

We explored the impact of different non-pharmaceutical control interventions and strategies (packages and sequences of interventions) that may effectively be implemented in African countries to mitigate COVID-19 epidemics. Short of an indefinite-duration lock-down, none of the interventions would likely avert very large epidemics that result in high mortality and extreme pressure on health services. However, both self-isolation of symptomatic people and moderate physical distancing could translate into relatively moderate but, in absolute terms, very sizable reductions in severe cases and deaths. The shielding option would likely require high levels of adherence and isolation to yield appreciable reductions in health service pressure, but could have a higher potential to reduce mortality in the short-term than other interventions, as it focuses on those who experience the highest CFR. This option also promotes herd immunity through mixing of other age groups and thus carries a lesser risk of further epidemic peaks once measures are lifted.

Different shielding arrangements could be considered, ranging from individual arrangements wherever people already live in multi-room houses or compounds, to neighbours or extended family-members grouping the most vulnerable individuals in vacated houses, to larger, albeit epidemiologically riskier re-housing (e.g. in quarantined street blocks). To avoid the problem of transmission within the shielded population, all such arrangements would need to eliminate any traffic of external people in and out of shielded accommodation as much as possible ensuring basic needs are met, while also instituting infection control barriers, e.g. a designated exchange point for supplies and safe social interactions and limiting contact within the shielded population. While our model does not explore the micro-level dynamics of how seeding of infection into these accommodations would affect residents, we showed, as expected, that the amount of contact among high-risk shielded people matters: zero contact, equivalent to individual shielding, equates to the highest effect of the intervention, while an increase of contact from baseline, e.g. if shielded people are rehoused in more crowded conditions than in their households of origin, dampens the utility of this approach and could even lead to an increase in cases compared to the baseline of no intervention.

We next combined the above interventions into a set of strategies, with a horizon of 12 months, that countries could consider. We assumed that any strategy would feature, at a minimum, self-isolation of symptomatic cases. Our predictions suggest that countrywide lockdowns of two months, if effective, would temporarily suppress and delay epidemics for around 2 months, as noted in Europe: this reprieve would potentially enable countries to mobilise resources and plan the implementation of the next phase of their strategy. These findings do not in themselves support lockdown measures as a universal solution, however: such measures may be ineffective (i.e. fail to achieve a contact rate reduction consistent with effective reproduction number < 1, the condition for suppression) or more harmful than beneficial, including in health terms, if they severely disrupt economies and livelihoods or encounter mistrust and community resistance. Rather, our predictions merely indicate that well-implemented lockdowns would achieve the intended effect.

If lockdowns are not implemented, or after they end, we predict that a combination of general physical distancing and shielding high-risk individuals could be a potentially achievable mitigation strategy for countries to consider. Physical distancing entails a difficult trade-off between reducing attack rates (and hence epidemic peak size) and extending the duration of the epidemic, which in turn increases the period over which shielding should be maintained (as individuals will require to be shielded until well after the epidemic peak has finished). While stringent physical distancing (e.g. 50% reduction in extra-household contacts) would have a large impact, such reductions may only be achieved through socio-economically damaging and potentially unacceptable restrictions to work, education and/or other forms of public life. By contrast, a 20% reduction in transmission may be more achievable and sustainable - in some settings, this could involve a combination of hygiene promotion, increased access to water, soap and other cleaning supplies (e.g. through state subsidies) and curtailment of some gatherings outside of work and school. It is unknown at present whether shielding is at all feasible and can attain our suggested target of 80% contact reduction between high- and low-risk people for 80% of high-risk people. Even at lower effectiveness levels, however, shielding would still offer benefits, particularly in terms of mortality, and accordingly need not be discounted as an option, particularly if it can be designed and led by communities themselves^33^, thereby requiring fewer resources than a top-down intervention.

We only show estimates for the first 12 months in our analysis to make short-term predictions of the impact of different intervention strategies. There are still many unknowns about how SARS-CoV-2 would behave in in different contexts in Africa (rural, urban, peri-urban, displacement settings, etc.) and how people will respond to policy measures; hence policy makers will continuously need to revisit the strategy to take current developments into account. Our analysis does not necessarily reflect the total epidemic size, as additional severe and critical cases could accrue during the second year, especially under strategies that focus heavily on physical distancing or if natural immunity to infection is short-lived.^4^

### Comparison with other studies

Although several studies have looked at the spread of COVID-19 in African countries^34–38^, we are only aware of one other modelling study which considers the impact of different interventions on the spread of COVID-19 in Africa. Walker et al.^39^ used a similar SEIR model and predicted a near 90% reduction in cases for sub-Saharan Africa assuming a 75% reduction in contacts starting at an incidence of 0.2 deaths per 100,000 population per week and sustained over the first 250 days of an epidemic. Comparisons between these studies are complicated due to different time periods of models and strategies investigated, but both point to a large unmitigated epidemic which can be reduced substantially due to strong physical distancing measures. However, even with these measures in place all models suggest a high burden of disease and mortality across Africa.

### Study limitations

While SEIR models have successfully been used to model COVID-19 epidemics to date in Europe, our predictions for African countries are subject to potential inaccuracy. Transmissibility may vary considerably across Africa, and it is possible that countries with very concentrated urban populations would see an acute exponential rise in cases, with a secondary, flatter curve affecting outlying rural regions: models accounting for these very distinct settlement types may be more useful for national planning. Consequently, the duration of time that countries will be affected by COVID-19 according to our model should be treated with caution. While we accounted for age distributions, we did not have country-specific data on contact patterns among age groups. Instead, we used synthetic contact matrices extrapolated from European data by using local data on household, workplace and school composition in the African settings considered. A sensitivity analysis with empirical African contact pattern data suggested lower effects of general distancing and higher effects of shielding, but these matrices were collected in specific areas and may not be representative of contact patterns within the entire country or indeed Africa as a whole.

Our results are also heavily affected by disease severity assumptions. We applied age-specific risks of developing severe disease per infection, as estimated using data from China and the Diamond Princess outbreak, but shifted these to earlier life by a decade to represent plausible differences in biological age in Africa resulting from life-course exposures. This crude approach may be confounded by differences in the age-specific prevalence of co-morbidities in African countries, as well as inter-country differences in comorbidity prevalence. Specifically, conditions with potential (tuberculosis^39^) and as yet undocumented (HIV, undernutrition, sickle-cell disease) interactions with SARS-CoV-2 infection are far more prevalent in Africa than China and affect relatively young age groups^10^: these could increase COVID-19 severity overall and shift the distribution of severe cases to younger age. Additionally, the proportion of detected and correctly managed cases of non-communicable diseases of known import to COVID-19 progression (cardiovascular disease, diabetes, chronic obstructive pulmonary disease, chronic kidney disease) is far lower in most of Africa than in China and Europe: this may further exacerbate disease severity. These differing patterns of co-morbidity may also affect the proportion of patients requiring intensive care and case-fatality: for the latter, we have provided illustrative findings assuming age-constant increases in Africa compared to China, based on an assumption of very limited treatment. These findings will need to be refined as more evidence accrues on the virus’ CFR in African settings.

Severity and CFR assumptions mostly affect the usefulness of the shielding option. Shielding criteria should include a diagnosis of co-morbidity, and as such our findings, based solely on an age criterion, are underestimates insofar as they exclude younger people with known comorbidities. However, in practice the low non-communicable disease treatment coverage in Africa means the age criterion would largely define who is shielded: lowering this threshold, e.g. to 50 years, would capture a larger fraction of undiagnosed co-morbidities. Tiered shielding approaches, whereby middle-aged moderate-risk people benefit from partial distancing measures (e.g. support to stay home from work), may also be worth considering.

### Conclusions

COVID-19 epidemics in African countries may bear very serious and multi-faceted impacts. Nevertheless, preventive strategies to substantially mitigate these impacts are not foreclosed to African governments and societies, particularly if they receive assistance from humanitarian and development actors, diaspora communities, faith-based institutions, the private sector and others within these societies who have means to assist the response. Self-isolation and moderate physical distancing can be effective interventions. The shielding option can be proactively explored to test locally appropriate solutions. As the epidemic progresses, real-time modelling and strategy evaluation should be made available to African countries that do not yet have this expertise: this requires coordination and proactive support from the worldwide scientific community, as well as close exchange of information between modelling teams and country-based surveillance, so as to gradually refine predictions.

## Data Availability

We used publicly available data.

## Supplementary material

### Model structure

We used a stochastic compartmental dynamic transmission model, stratified into age groups *i*. The model’s structure is depicted in Figure S1.

**Supplemental Figure S1.**
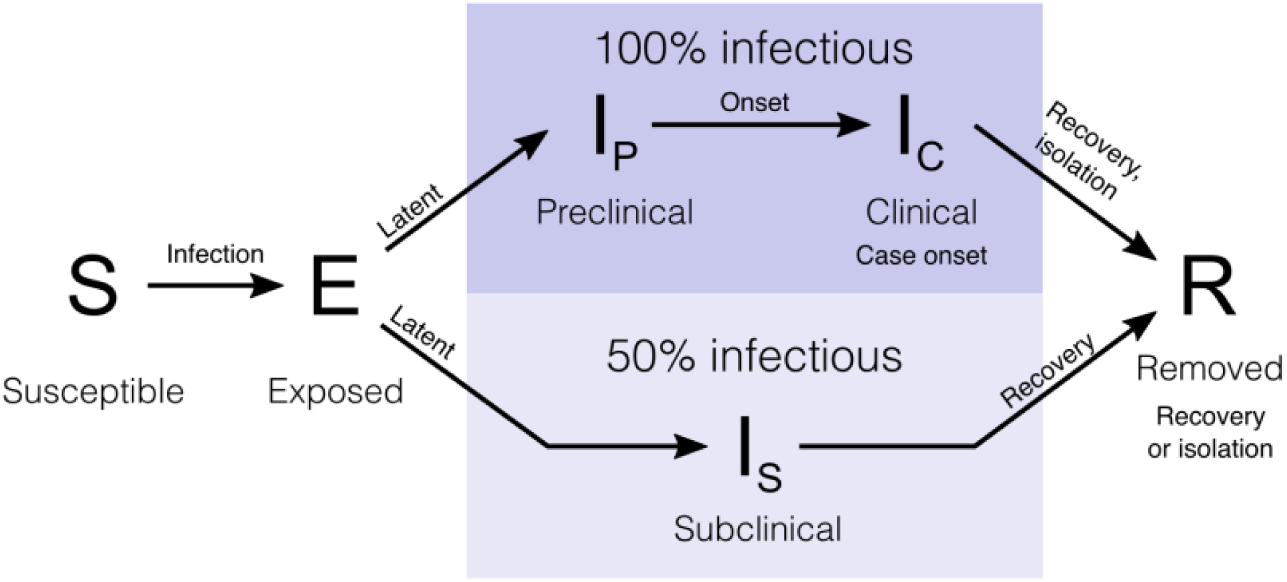
State transitions in the model. Individuals in the stochastic compartmental model are classified into susceptible, exposed (infected but not yet infectious), infectious (preclinical, clinical, or subclinical), and recovered states. The model is stratified into 5-year age bands. Taken from Davies et al.^21^

The model tracks a country’s population over discrete 6-hour increments. Within every age group, the population is distributed into compartments of susceptible individuals (*S*), who become exposed (*E*) after effective contact with an infectious person (exposed here means infected but not yet infectious). At the end of the latency period, people divide into clinical (symptomatic) and sub-clinical (asymptomatic) cases with complementary probabilities *y_i_* and *1* − *y_i,_* at which point infectiousness starts. Clinical cases experience a pre-clinical but infectious state *I_P_* followed by a clinical, infectious state *I*_c_. Sub-clinical cases (*I_s_*) are assumed to be half as infectious as clinical cases. All individuals have the same duration of infectiousness. The clinical severity of cases is not assumed to affect infectiousness. Individuals either recover or die, transitioning to the removed (*R*) compartment. Transitions to different clinical states (symptom onset to severe case, critical or not; from onset of severe symptoms to recovery or death) occur with delays drawn from the literature: these delays have no bearing on the force of infection / transmissibility.

### Force of infection and transmissibility

The force of infection is defined as the rate at which susceptible people enter the exposed compartment, and is computed for any age group *i* and time increment *t* as

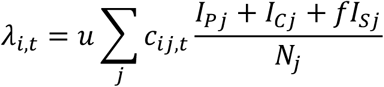

where *u* is the probability of infection per contact with an infectious person, *c_ij_* is the number of contacts that an individual in age group *i* has with individuals in age group *j* per time increment (drawn from the contact matrix), and 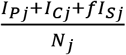 is the probability that any age j individual contacted is in fact infectious, with *f* denoting the relative infectiousness of subclinical cases, compared to clinical cases.

The amount of time a given individual spends in states *E, I_P_, l_c_*, or *I_s_* is drawn from distributions *d_E_, d_P_, d_c_* and *d_s_*, respectively (Table S1). The basic reproduction number *R*_0_ is defined as the average number of secondary infections generated by a typical infectious individual in a fully susceptible population and is calculated as the absolute value of the dominant eigenvalue of the next generation matrix (NGM), which was derived by linearizing the system at epidemic equilibrium.^40^ Lastly, *u* is derived for any stochastic run from the ratio of this eigenvalue and the *R*_0_ value selected for that run.

### Parameter values

The model was implemented stochastically by selecting random values of parameters from their uncertainty distributions (Table S1).

### Transmission dynamics and state transition parameters

**Table S1.** Model parameters relevant to transmission and state transitions. Adapted from Davies et al.^21^.

| Parameter | Description | Value | Reference |
| --- | --- | --- | --- |
| $d_E$ | Latent period in days | $\sim \text{gamma}(\mu = 4, k = 4)$ | 41–43 |
| $d_p$ | Duration of preclinical infectiousness in days | $\sim \text{gamma}(\mu = 1.5, k = 4)$ | 44 |
| $d_c$ | Duration of clinical infectiousness in days | $\sim \text{gamma}(\mu = 3.5, k = 4)$ | 41–43 |
| $d_s$ | Duration of subclinical infectiousness in days | $\sim \text{gamma}(\mu = 5, k = 4)$ | Assumed to be the same as duration of total infectious period for clinical cases |
| $u$ | Probability of transmission per contact with an infectious individual | See text | Derived |
| $y_i$ | Probability of clinical symptoms on infection for age group $i$ | Age-dependent, as estimated in <sup>22</sup> | 22 |
| $f$ | Relative infectiousness of subclinical cases | 50% | Assumed |
| $c_{ij}$ | Number of age-$j$ individuals contacted by an age-$i$ individual per day | Country-specific synthetic contact matrix | 24 |
| $R_0$ | Basic reproduction number | $\sim \text{normal}(\mu = 2.6, s = 0.5)$ | 25 |
| $N_i$ | Number of age $i$ individuals | Demographic estimates | 23 |
| $\Delta t$ | Time step for discrete-time simulation | 0.25 days | |
| | Delay from symptom onset to becoming a severe case in days | $\sim \text{gamma}(\mu = 7, k = 7)$ | 45,46 |
| | Duration of severe, non-critical disease in days | $\sim \text{gamma}(\mu = 8, k = 8)$ | Duration based on NHS data for J12: viral pneumonia, not elsewhere classified <sup>47</sup> |
| | Duration of severe, critical disease in days | $\sim \text{gamma}(\mu = 10, k = 10)$ | <sup>46</sup> |
|  | Proportion of hospitalised cases that require critical care | 30% | <sup>46</sup> |
| | Delay from symptom onset to death in days | $\sim \text{gamma}(\mu = 22, k = 22)$ | <sup>45,48</sup> |

### Severity parameters

We used the same severity estimates as in Davies et al^21^: these reflect the early COVID-19 outbreak in China^48^, corrected based on data from the Diamond Princess cruise ship outbreak.^49^ However, in African and low-income countries, someone’s vulnerability to infection may correspond to that of an individual with greater chronological age in a high-income setting due to life-course effects like malnutrition, infections and often unmanaged non-communicable diseases. It is not yet known whether this increased vulnerability will affect COVID-19 age-specific disease risk in African populations. To provide conservative estimates, we shifted age-specific severity risks by 10 years towards younger ages. In addition, we multiplied the CFRs of severe and critical cases by a factor of 1.5 to account for lower access to care. Age-specific severity estimates used are presented in Table S2.

**Table S2.** Model parameters relevant to age-specific disease outcomes.

| Age | Risk of becoming a severe case | Proportion of severe cases that are critical | Case-fatality ratio among severe, non-critical cases | Case-fatality ratio among critical cases |
| --- | --- | --- | --- | --- |
| < 10 | 0.76% | 30% | 0.14% | 75% |
| 10 to 19 | 0.81% | 30% | 0.15% | 75% |
| 20 to 29 | 0.99% | 30% | 0.18% | 75% |
| 30 to 39 | 1.85% | 30% | 0.35% | 75% |
| 40 to 49 | 5.43% | 30% | 1.01% | 75% |
| 50 to 59 | 15.05% | 30% | 2.81% | 75% |
| 60 to 69 | 33.29% | 30% | 6.21% | 75% |
| 70+ | 61.76% | 30% | 11.52% | 75% |

## Additional analyses

**Supplemental Figure S2.**
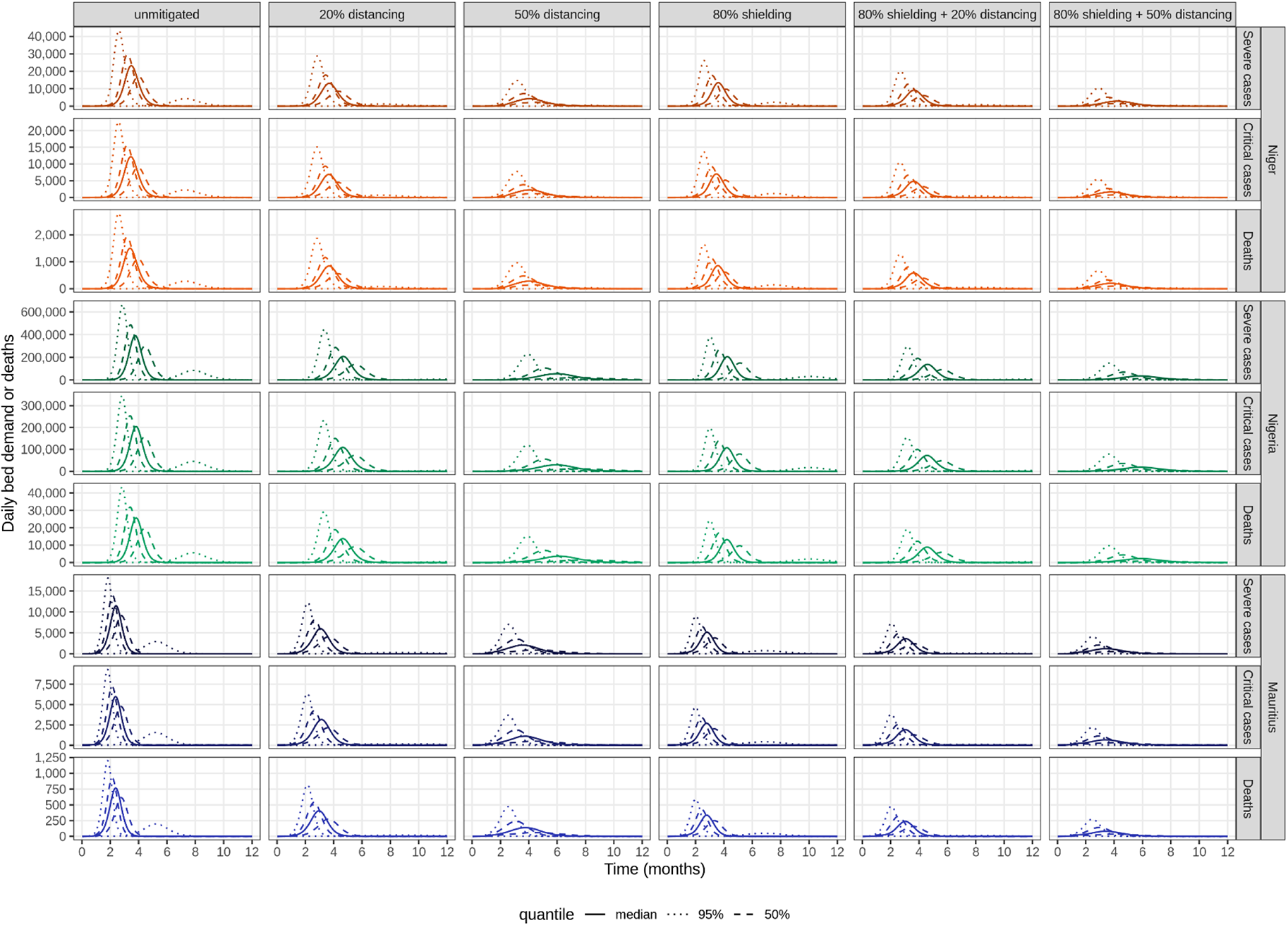
Bed demand and deaths during the first 12 months of the epidemic, under different strategies. We show runs representing the median, 95%, and 50% quantiles of total number of the corresponding outcome, after 12 months.

**Supplemental Figure S3.**
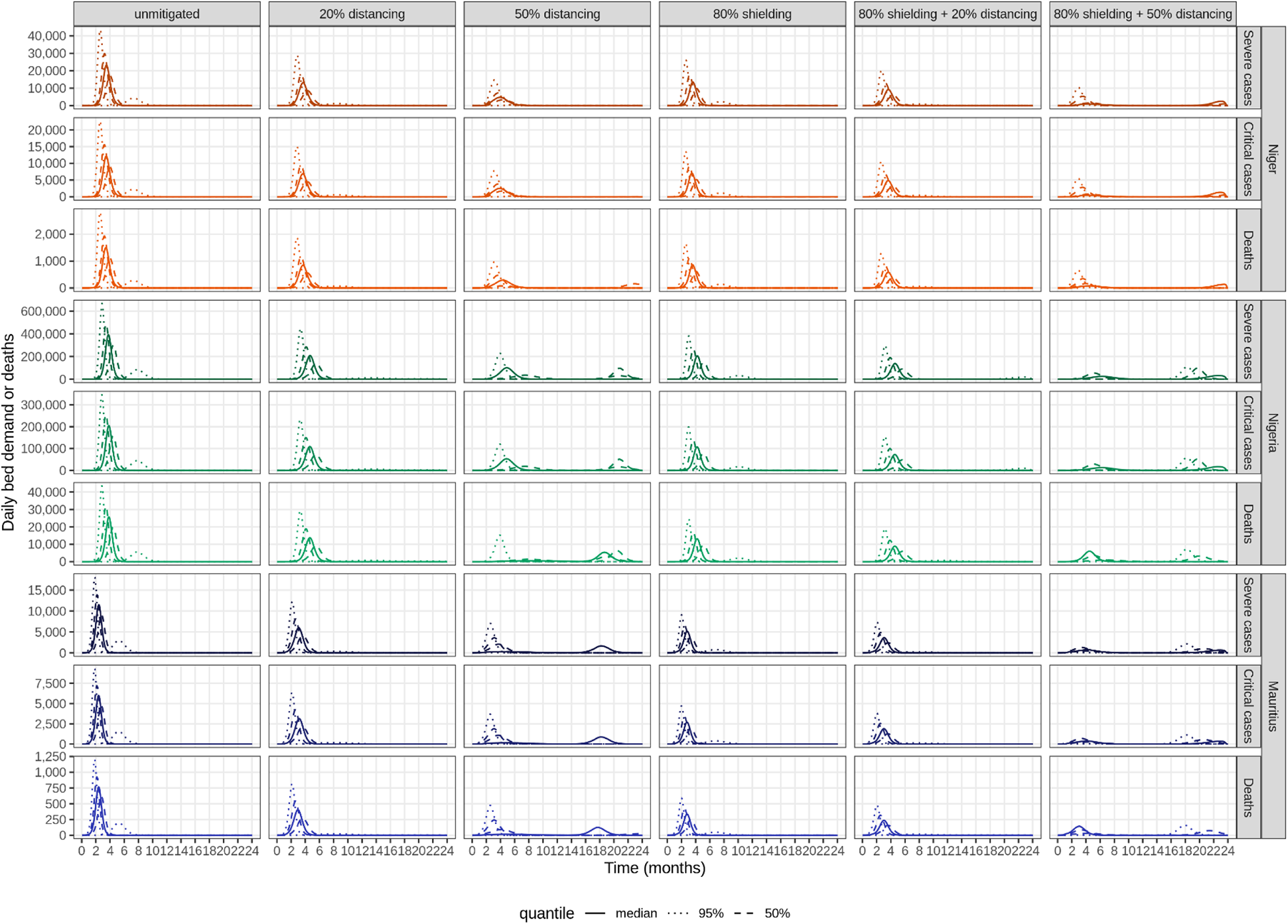
Bed demand and deaths during the first 24 months of the epidemic, under different strategies. All interventions are stopped after 12 months, and no new interventions are assumed to be in place in the second year. We show runs representing the median, 95%, and 50% quantiles of total number of the corresponding outcome, after 24 months.

**Supplemental Figure S4.**
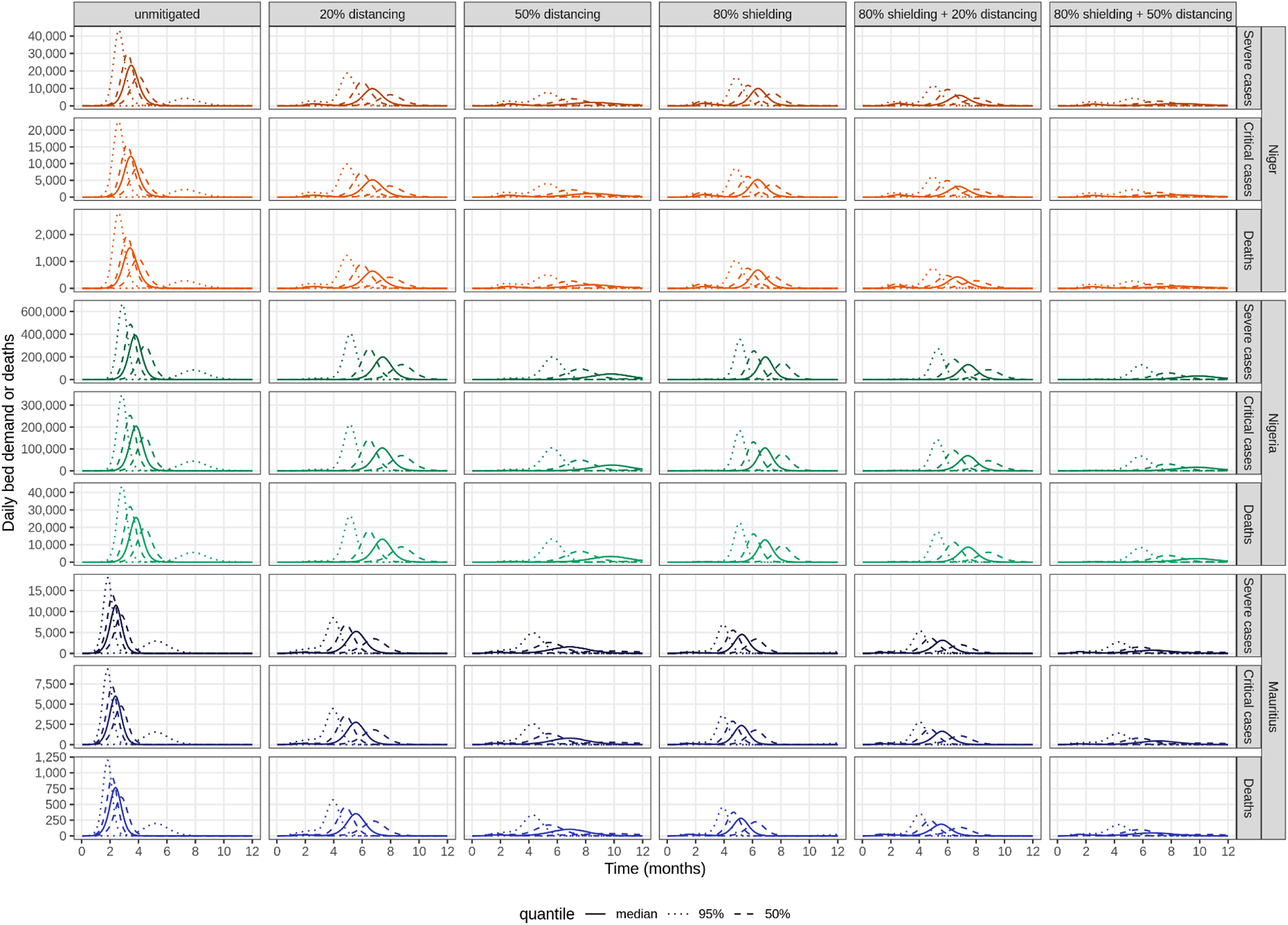
Bed demand and deaths during the first 12 months of the epidemic, under different strategies preceded by a two-month lockdown. We show runs representing the median, 95%, and 50% quantiles of total number of the corresponding outcome, after 12 months.

Thick solid lines show the runs representing the median daily number of hospital beds needed for severe cases, critical cases, and the number of deaths during the first 12 months of the epidemic. Dashed lines show runs representing 50% quantiles, while dotted lines show runs representing 95% quantiles, of each respective value over 500 model runs. Except for the unmitigated scenario, all scenarios assume 50% self-isolation during the symptomatic period of all clinical cases throughout the entire course of the epidemic and are preceded by a two-month lockdown where contacts outside of the household are reduced by 80%. Lockdown starts when daily incidence of symptomatic cases reaches 1 case per 10 000 people, while other strategies start once the lockdown is lifted. Non-lockdown distancing strategies assume 20% or 50% reduction in all contacts outside of the household. Shielding strategies assume shielding of 80% of the population aged 60+, irrespective of underlying comorbidities, with an 80% reduction in contacts between the shielded and unshielded population, and no change in contacts within the shielded population.

**Supplemental Table S3.** Key outcomes over 12 months of strategies preceded by two-month lockdown.

**Niger**
| Key outcome | Unmitigated | 20% distancing | 50% distancing | 80% shielding | 80% shielding + 20% distancing | 80% shielding + 50% distancing |
| --- | --- | --- | --- | --- | --- | --- |
| Symptomatic cases | 4,101,000<br>(1,873,000 - 5,240,000) | 2,771,000<br>(163,000 - 3,990,000) | 1,311,000<br>(85,000 - 2,711,000) | 3,326,000<br>(206,000 - 4,383,000) | 2,667,000<br>(153,000 - 3,813,000) | 1,224,000<br>(97,000 - 2,559,000) |
| Severe cases | 106,000<br>(39,000 - 154,000) | 68,000<br>(3,000 - 110,000) | 33,000<br>(2,000 - 76,000) | 60,000<br>(4,000 - 89,000) | 48,000<br>(3,000 - 76,000) | 22,000<br>(2,000 - 51,000) |
| Critical cases | 45,000<br>(17,000 - 66,000) | 29,000<br>(1,000 - 47,000) | 14,000<br>(1,000 - 33,000) | 26,000<br>(2,000 - 38,000) | 21,000<br>(1,000 - 33,000) | 10,000<br>(1,000 - 22,000) |
| Deaths | 39,000<br>(14,000 - 56,000) | 25,000<br>(1,000 - 40,000) | 12,000<br>(1,000 - 28,000) | 21,000<br>(1,000 - 31,000) | 17,000<br>(1,000 - 27,000) | 8,000<br>(1,000 - 18,000) |
| Symptomatic attack rate | 16.9%<br>(7.7 - 21.6) | 11.4%<br>(0.7 - 16.5) | 5.4%<br>(0.4 - 11.2) | 13.7%<br>(0.9 - 18.1) | 11%<br>(0.6 - 15.8) | 5.1%<br>(0.4 - 10.6) |
| Epidemic peak (months) | 3 (3 - 7) | 7 (5 - 11) | 7 (3 - 11) | 6 (5 - 11) | 7 (5 - 11) | 6 (2 - 10) |
| Peak deaths | 1,500<br>(300 - 2,800) | 600<br>(0 - 1,300) | 100<br>(0 - 500) | 700<br>(0 - 1,200) | 400<br>(0 - 800) | 100<br>(0 - 300) |
| Peak non-ICU beds needed | 23,200<br>(4,400 - 43,200) | 9,900<br>(500 - 19,800) | 2,000<br>(400 - 8,200) | 10,600<br>(700 - 18,100) | 6,600<br>(500 - 13,000) | 1,300<br>(400 - 5,100) |
| Peak ICU beds needed | 12,200<br>(2,300 - 22,500) | 5,200<br>(300 - 10,400) | 1,100<br>(200 - 4,300) | 5,600<br>(300 - 9,500) | 3,500<br>(300 - 6,800) | 700<br>(200 - 2,700) |

**Nigeria** **\* THIS STUDY HAS NOT YET BEEN PEER-REVIEWED \***
| Key outcome | Unmitigated | 20% distancing | 50% distancing | 80% shielding | 80% shielding + 20% distancing | 80% shielding + 50% distancing |
| --- | --- | --- | --- | --- | --- | --- |
| Symptomatic cases | 48,728,000<br>(27,753,000 - 56,537,000) | 37,553,000<br>(124,000 - 48,968,000) | 18,807,000<br>(110,000 - 37,679,000) | 42,207,000<br>(229,000 - 50,862,000) | 36,099,000<br>(158,000 - 46,849,000) | 17,859,000<br>(138,000 - 35,787,000) |
| Severe cases | 1,631,000<br>(734,000 - 2,119,000) | 1,179,000<br>(3,000 - 1,733,000) | 579,000<br>(3,000 - 1,323,000) | 952,000<br>(4,000 - 1,300,000) | 789,000<br>(3,000 - 1,165,000) | 371,000<br>(3,000 - 858,000) |
| Critical cases | 699,000<br>(314,000 - 908,000) | 505,000<br>(1,000 - 743,000) | 248,000<br>(1,000 - 567,000) | 408,000<br>(2,000 - 557,000) | 338,000<br>(1,000 - 499,000) | 159,000<br>(1,000 - 368,000) |
| Deaths | 605,000<br>(271,000 - 790,000) | 438,000<br>(1,000 - 646,000) | 217,000<br>(1,000 - 495,000) | 340,000<br>(2,000 - 468,000) | 283,000<br>(1,000 - 419,000) | 133,000<br>(1,000 - 309,000) |
| Symptomatic attack rate | 23.6%<br>(13.5 - 27.4) | 18.2%<br>(0.1 - 23.8) | 9.1%<br>(0.1 - 18.3) | 20.5%<br>(0.1 - 24.7) | 17.5%<br>(0.1 - 22.7) | 8.7%<br>(0.1 - 17.4) |
| Epidemic peak (months) | 4 (3 - 8) | 7 (5 - 12) | 8 (4 - 12) | 7 (5 - 12) | 7 (5 - 12) | 8 (3 - 12) |
| Peak deaths | 25,600<br>(5,700 - 43,200) | 13,200<br>(0 - 26,700) | 3,300<br>(0 - 13,300) | 12,800<br>(100 - 22,200) | 8,500<br>(0 - 17,100) | 2,000<br>(0 - 8,100) |
| Peak non-ICU beds needed | 391,000<br>(85,800 - 662,900) | 199,400<br>(500 - 405,800) | 49,800<br>(500 - 199,900) | 199,400<br>(800 - 347,400) | 131,400<br>(600 - 266,000) | 31,200<br>(600 - 125,600) |
| Peak ICU beds needed | 204,500<br>(45,300 - 343,900) | 104,900<br>(300 - 212,100) | 26,300<br>(300 - 105,300) | 104,600<br>(400 - 180,900) | 69,100<br>(300 - 139,100) | 16,500<br>(300 - 66,100) |

**Mauritius**
| Key outcome | Unmitigated | 20% distancing | 50% distancing | 80% shielding | 80% shielding + 20% distancing | 80% shielding + 50% distancing |
| --- | --- | --- | --- | --- | --- | --- |
| Symptomatic cases | 490,000<br>(314,000 - 546,000) | 389,000<br>(6,000 - 478,000) | 244,000<br>(5,000 - 392,000) | 392,000<br>(34,000 - 448,000) | 345,000<br>(7,000 - 419,000) | 210,000<br>(6,000 - 340,000) |
| Severe cases | 44,000<br>(24,000 - 53,000) | 32,000<br>(0 - 43,000) | 20,000<br>(0 - 35,000) | 23,000<br>(2,000 - 30,000) | 20,000<br>(0 - 27,000) | 12,000<br>(0 - 21,000) |
| Critical cases | 19,000<br>(10,000 - 23,000) | 14,000<br>(0 - 19,000) | 9,000<br>(0 - 15,000) | 10,000<br>(1,000 - 13,000) | 9,000<br>(0 - 11,000) | 5,000<br>(0 - 9,000) |
| Deaths | 17,000<br>(9,000 - 21,000) | 12,000<br>(0 - 17,000) | 8,000<br>(0 - 13,000) | 9,000<br>(1,000 - 11,000) | 7,000<br>(0 - 10,000) | 4,000<br>(0 - 8,000) |
| Symptomatic attack rate | 38.5%<br>(24.7 - 42.9) | 30.6%<br>(0.5 - 37.6) | 19.2%<br>(0.4 - 30.8) | 30.8%<br>(2.7 - 35.2) | 27.1%<br>(0.6 - 32.9) | 16.5%<br>(0.5 - 26.7) |
| Epidemic peak (months) | 2 (2 - 5) | 5 (4 - 11) | 6 (2 - 12) | 5 (4 - 10) | 5 (3 - 11) | 6 (2 - 11) |
| Peak deaths | 800<br>(200 - 1,200) | 300<br>(0 - 600) | 100<br>(0 - 300) | 300<br>(0 - 400) | 200<br>(0 - 300) | 100<br>(0 - 200) |
| Peak non-ICU beds needed | 11,500<br>(3,000 - 18,000) | 5,200<br>(100 - 8,800) | 1,500<br>(100 - 4,600) | 4,500<br>(400 - 6,500) | 3,000<br>(100 - 5,000) | 800<br>(100 - 2,500) |
| Peak ICU beds needed | 6,000<br>(1,600 - 9,300) | 2,700<br>(0 - 4,600) | 800<br>(0 - 2,400) | 2,400<br>(200 - 3,400) | 1,600<br>(0 - 2,600) | 400<br>(0 - 1,300) |

\* THIS STUDY HAS NOT YET BEEN PEER-REVIEWED \*

**Supplemental Figure S5.**
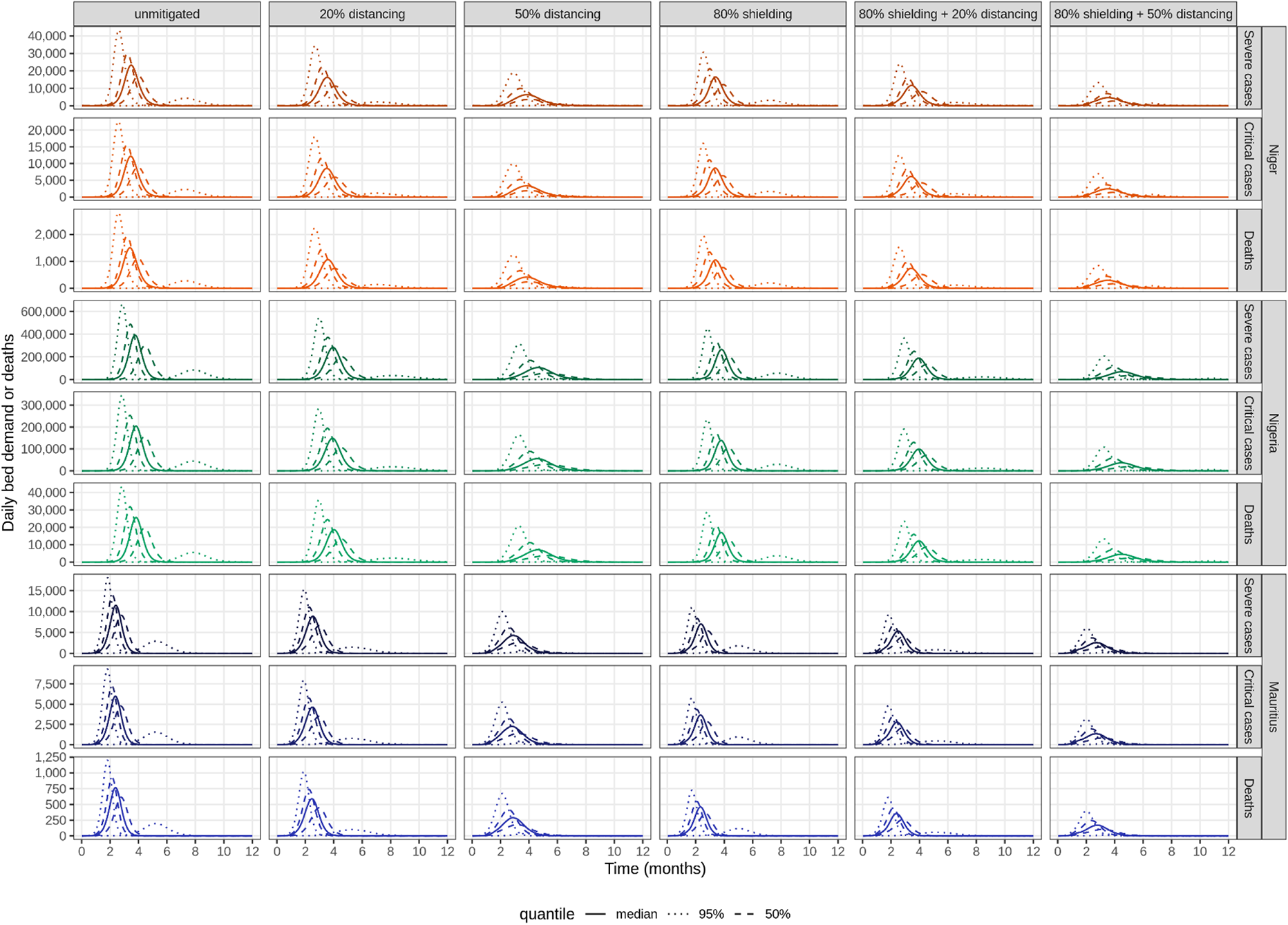
Sensitivity analysis: bed demand and deaths during the first 12 months of the epidemic, under different strategies but in the absence of self-isolation. We show runs representing the median, 95%, and 50% quantiles of total number of the corresponding outcome, after 12 months.

**Supplementary Table S4.**
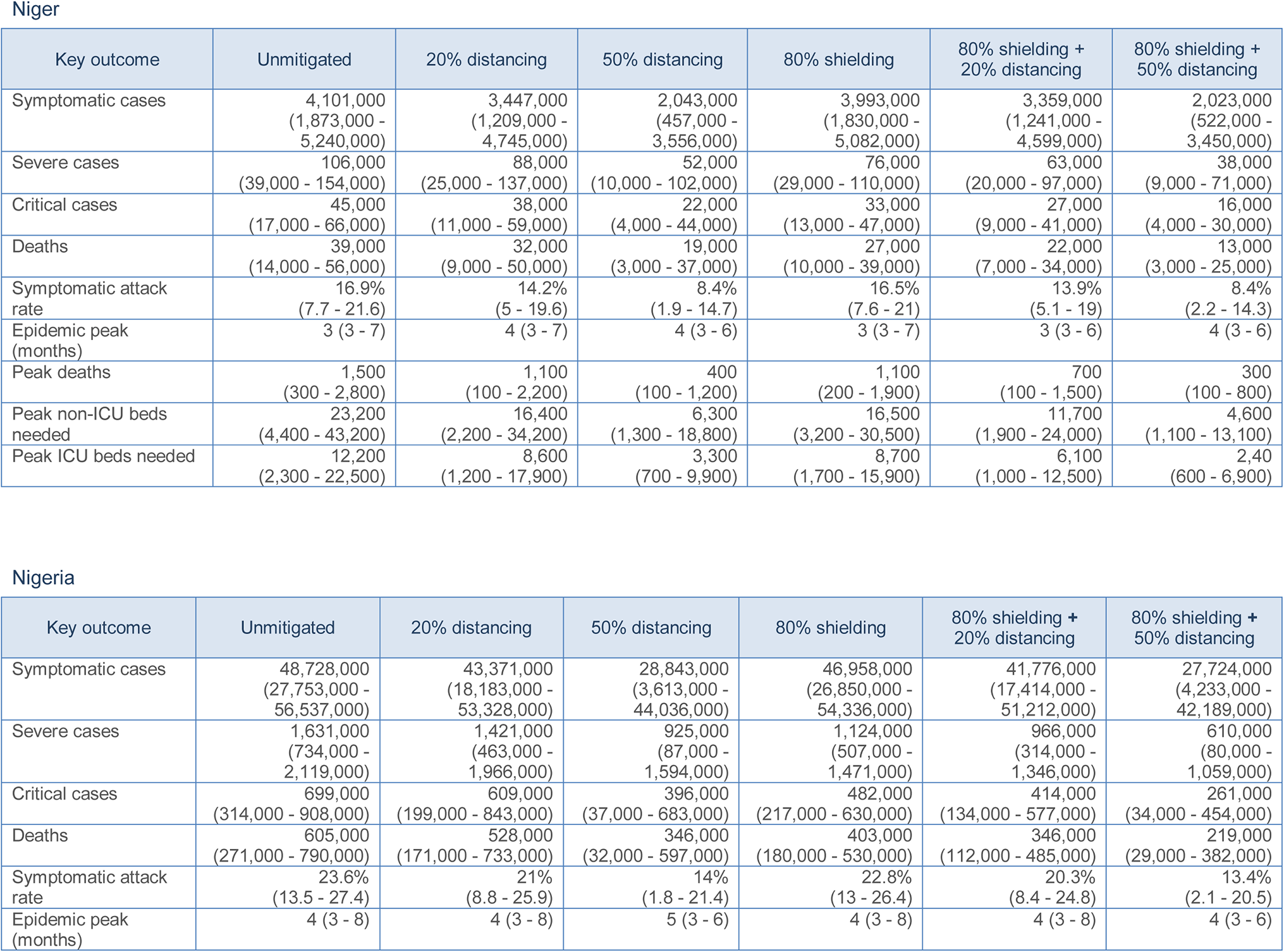
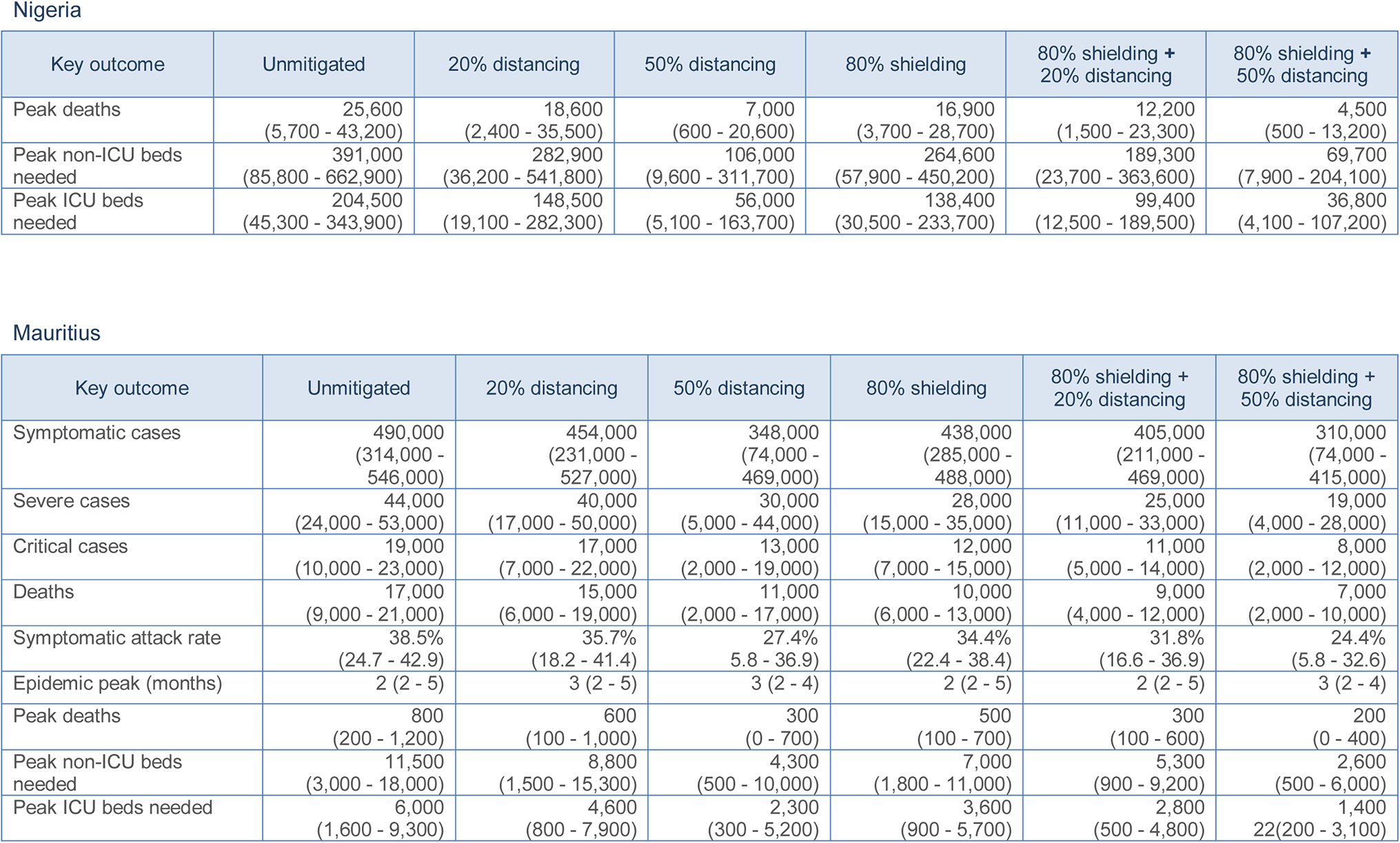
Sensitivity analysis: key outcomes during the first 12 months of the epidemic, under different strategies but in the absence of self-isolation.

## Estimated impact in countries with empirical contact data: Kenya, Uganda, and Zimbabwe

We replicated the individual intervention analysis in three countries for which we had empirical contact matrices (Kenya, Uganda and Zimbabwe). As these data came from household sample surveys, we took one new bootstrapped sample from the contact matrix in each model run.

Unmitigated epidemic projections are shown in Figure S6. Figure S7 shows the effects of self-isolation and physical distancing, which have been run for a subset of scenarios as used in the main paper for Niger, Nigeria and Mauritius only. Effect sizes appear lower than for the main analysis of Niger, Nigeria and Mauritius using synthetic contact matrices.

Lastly, as shown in Figure S8, the potential effect of shielding appears about 10% higher than for the main analysis, but with wider uncertainty due to the propagation of contact matrix sampling error. In addition, Uganda, Kenya, and Zimbabwe have different age-distributions, so results are not directly comparable. This analysis shows shielding to be comparatively more sensitive to changes in contact among shielded individuals, with a substantial loss of effect when shielded people greatly increase their contacts, e.g. if they are rehoused in overcrowded conditions.

**Supplemental Figure S6.**
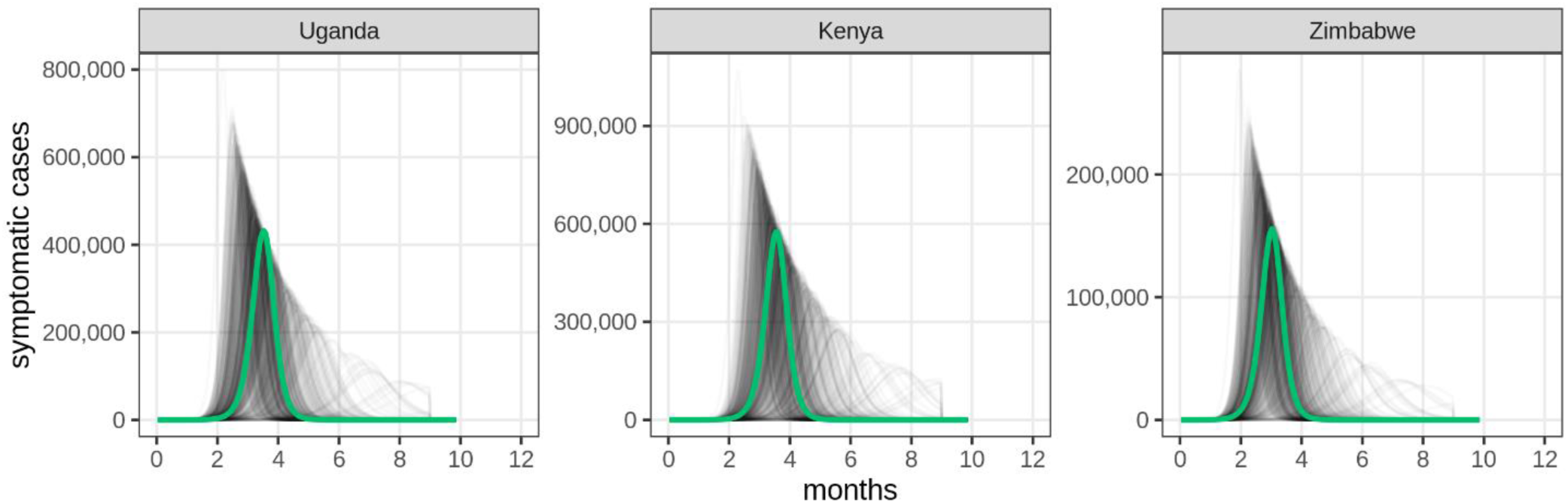
Projected incidence of symptomatic COVID-19 cases over time for simulations of an unmitigated epidemic, by country with empirical contact matrix data. The green line shows the run corresponding to the median total of cases at the end of the simulation across all model runs. Grey lines show individual stochastic model runs, where R0 in each run was sampled from a normal distribution with mean 2.6 and standard deviation 0.5.

**Supplementary Figure S7:**
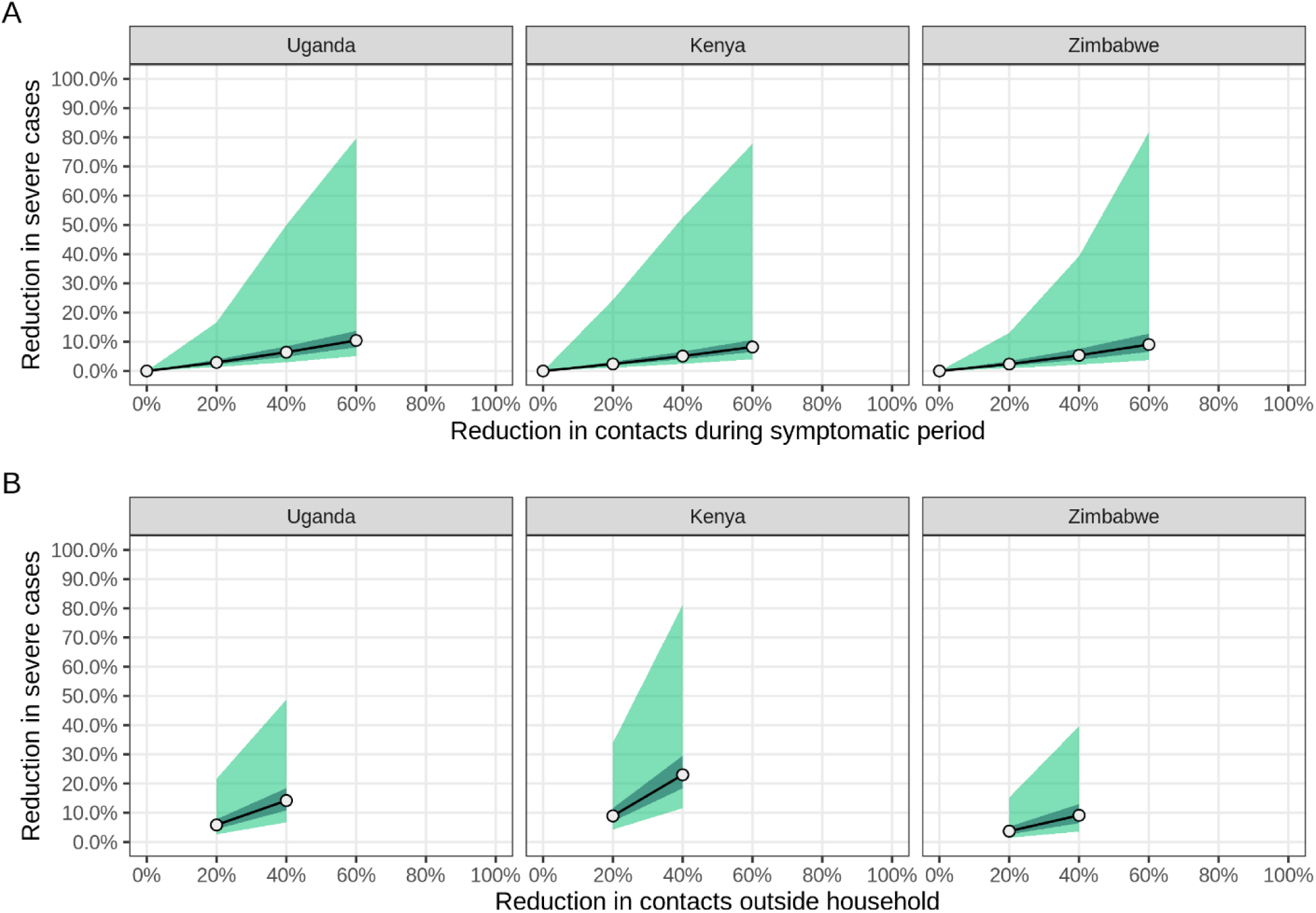
Estimated reduction in severe cases following A) self-isolation of symptomatic individuals and B) population-wide physical distancing, by country with empirical contact matrix data. Medians (circles), 95% (light-green area) and 50% (dark green area) quantiles for the percentage reduction in severe cases (patients requiring hospitalisation) during the first 12 months of the epidemic for different levels of compliance, for each country, across 500 model runs.

**Supplementary Figure 8:**
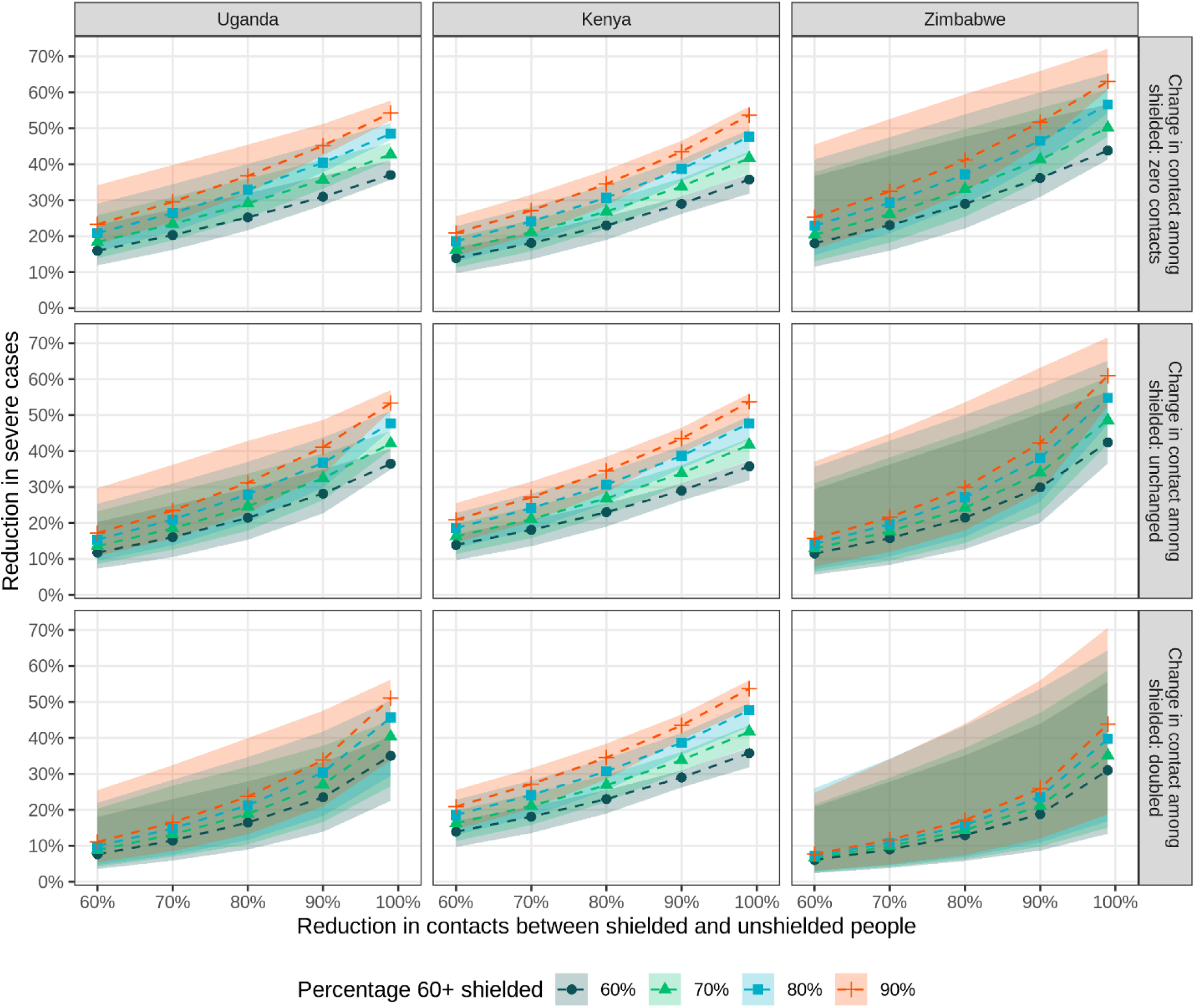
Estimated reduction in severe cases following shielding of high-risk individuals, by country with empirical contact matrix data. Medians (dashed lines) and 95% quantiles (shaded areas) of the percentage reduction in severe cases during the first 12 months of the epidemic for different levels of reduction in contacts between shielded and unshielded people (x axis), different level of contacts among shielded people (facet rows), and for different percentages of ≥ 60 years old being shielded (see legend) for each country, across 500 model runs.

### Sensitivity to transmissibility assumption (*R*_0_)

Uncertainty intervals in our results were mainly reflective of the range of *R*_0_ values we sampled in the model. Here we show our results stratified by *R*_0_ values, as sampled across 500 runs from a normal *R*_0_ distribution with mean 2.6 and standard deviation of 0.5.

Supplemental Figure S9 shows the unmitigated epidemic in each country for every model run (i.e. randomly sampled *R*_0_ value) where *R*_0_ is grouped into values <2, between 2 and 3, and >3. Epidemics with high *R*_0_ have a higher peak number of cases and total number of cases, and will peak earlier, whereas epidemics with a low *R*_0_ will have a lower peak number of cases and total number of cases, and will peak later.

**Supplemental Figure S9.**
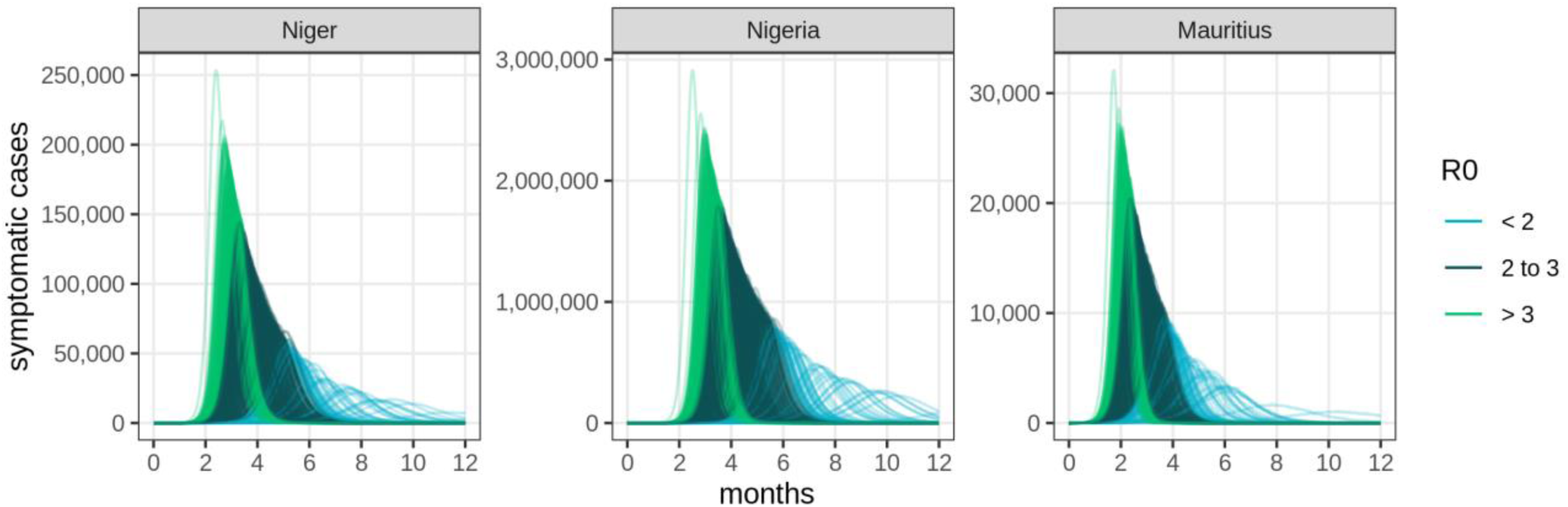
**Projected incidence of symptomatic COVID-19 cases over time for simulations of an unmitigated epidemic, by** *R*_0_ value.

Supplemental Figure S10A shows the impact of self-isolation and physical distancing (equivalent to Figure 2 in the main text), but for every single run and stratified by *R*_0_. High reductions as a result of self-isolation may only be achieved with very low values of *R*_0_, and are less likely in Niger, which has a younger population (i.e. symptomatic individuals are relatively fewer and thus contribute less to transmissibility). However, with higher values of *R*_0_, the impact of self-isolation is roughly similar across countries. Runs with smaller *R*_0_ are more affected by stochasticity of the model, which is why these estimates do not always continue their upward trend.

Supplemental Figure S10B shows the impact of physical distancing on the total number of severe cases. The impact of physical distancing can be substantially lower when *R*_0_ is higher, as greater reductions in contacts will be needed to bring *R* closer to 1.

**Supplemental Figure S10.**
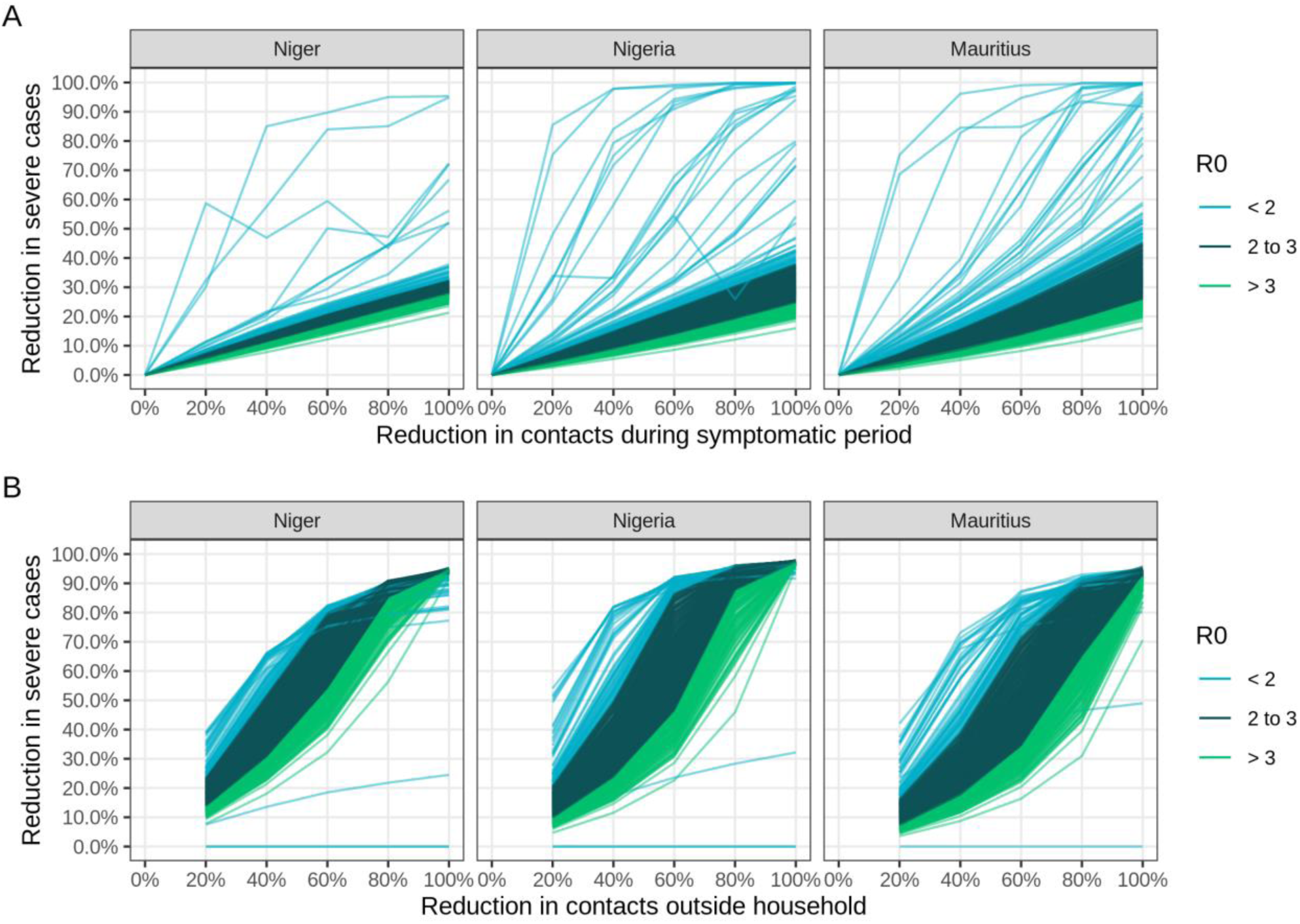
Impact of A. self-isolation and B. physical distancing, by *R*_0_ value sampled.

Lastly, Supplemental Figure S11 shows the impact of shielding 80% of the high-risk population under various reductions in contacts between the shielded and unshielded population, and changes in contact within the shielded population, for every single run. It is equivalent to Figure 3 in the main text. As shielding does not significantly affect the overall transmission dynamics in the population, impact does not vary much between different values of *R*_0_. Relative reductions could be substantially higher or lower at low values of *R*_0_, but reflect only a small absolute difference.

In Mauritius, where 18% of the population are 60+ years old (i.e. 15% are shielded under a shielding coverage of 80%), variation in *R*_0_ matters if contact within the shielded population changes from baseline, as this does significantly affect transmission dynamics in the overall population (the 15% of people shielded are relatively more infectious than the average).

**Supplemental Figure S11.**
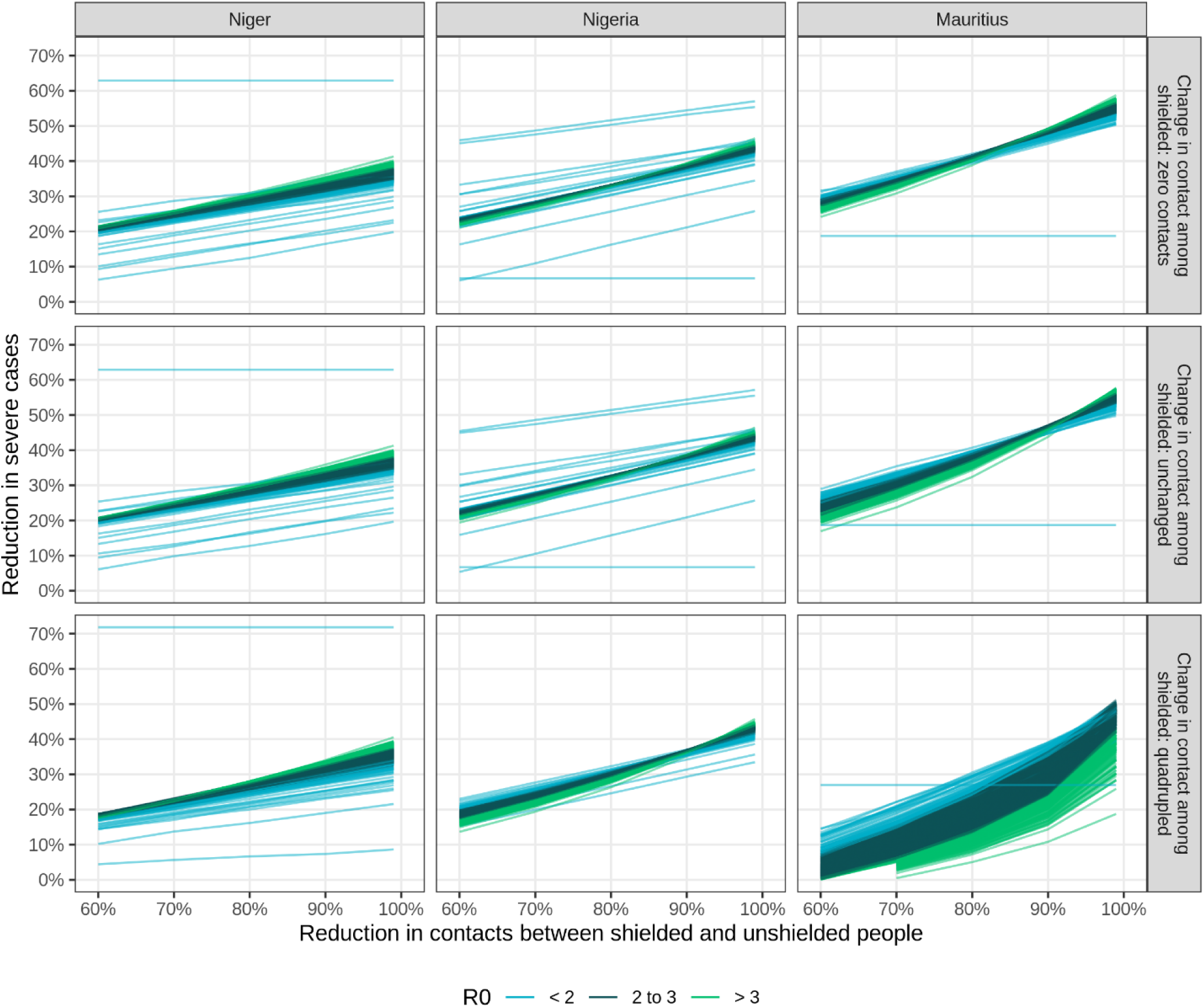
Impact of shielding by *R*_0_ value sampled.

